# National trends, demographic disparities, and phenotypic shifts in encephalopathy-associated mortality in the United States: A CDC WONDER analysis, 1999-2024

**DOI:** 10.64898/2026.09.21.26363584

**Authors:** Khaled Hamed, Ruweida Mohamed Bundid, Maha Ali Sayah

## Abstract

Encephalopathy is a lethal manifestation of global cerebral dysfunction spanning hypoxic, metabolic, toxic, and vascular etiologies, yet its population burden is difficult to quantify because it predominantly appears as a contributing rather than underlying cause of death. Here, we analyzed US multiple-cause death records from 1999 through 2024 using harmonized CDC WONDER datasets. A longitudinally consistent core definition of 10 ICD-10 codes identified 953,846 encephalopathy-associated deaths. The national age-adjusted mortality rate increased 28.4%, from 9.143 per 100,000 in 1999 to 11.738 in 2024, after peaking at 14.391 in 2021. Segmented log-linear regression identified four temporal phases: 1999-2004 (annual percent change [APC],-1.76%), 2004-2018 (APC, 2.57%), 2018-2021 (APC, 6.54%), and 2021-2024 (APC, −5.98%). Encephalopathy was listed as the underlying cause in only 21.6% of deaths in 2024. Unspecified encephalopathy surged more than 3.6-fold, while anoxic brain injury remained stable. Mortality remained higher in males than females (13.973 versus 9.778 per 100,000), and rates among non-Hispanic Black individuals were 1.90-fold those of non-Hispanic White individuals. Geographically, mortality clustered in the Midwest and South (13.144 and 13.120 per 100,000 in 2024), with persistent state-level dispersion across the study period. COVID-19 accounted for 10.9% of associated deaths in 2021 before falling to 1.4% in 2024. These findings reveal a nonlinear trajectory shaped by diagnostic shifts, pandemic disruption, and persistent demographic and geographic disparities.

## Introduction

Encephalopathy is not a single disease but a clinical-pathophysiologic state of global cerebral dysfunction that may arise from hypoxia, toxins, metabolic disturbance, vascular injury, infection, nutritional deficiency, or systemic critical illness.[1,2] Specific entities such as Wernicke encephalopathy, toxic-metabolic encephalopathy, hypertensive encephalopathy, and neonatal hypoxic-ischemic encephalopathy differ substantially in mechanism, age distribution, reversibility, and treatment.[1–6] This heterogeneity complicates surveillance because no single diagnosis code captures the syndrome and encephalopathy may be recorded as either an underlying or contributing cause of death.

Surveillance based only on the underlying cause can therefore miss deaths in which encephalopathy is documented as a contributing condition. The US National Vital Statistics System records both the underlying cause and multiple contributing causes on death certificates, enabling a broader view of mortality phenotypes.[7,8] Death-certificate analyses nevertheless require caution: diagnostic and certification practices can change over time, nonspecific codes may reflect documentation behavior as well as disease burden, race and ethnicity can be misclassified, and contributing causes indicate association with death rather than causal responsibility.[9,10] These issues are especially relevant to syndrome-level surveillance.

Recent US mortality studies using CDC WONDER have generally focused on single diagnoses or narrowly defined combinations and described temporal, demographic, and geographic variation.[11,12] For a heterogeneous syndrome such as encephalopathy, phenotype validation is additionally important: a single-code definition can mistake coding migration for epidemiologic change, whereas an excessively broad definition can introduce unrelated conditions. The overlap between the bridged-race and single-race mortality datasets in 2018-2020 also permits direct assessment of comparability across the database transition.

We therefore conducted a serial cross-sectional analysis of US multiple-cause mortality from 1999 through 2024. We prespecified a time-stable 10-code core phenotype (C10) as the primary definition, a comprehensive C11 phenotype including neonatal hypoxic-ischemic encephalopathy from 2006 onward, a narrow G93.4 definition, and broader sensitivity definitions. We examined long-term encephalopathy-associated mortality, tested robustness across nested phenotype definitions and cause-of-death position, characterized demographic and geographic disparities, and evaluated shifts in underlying etiologies, including during the COVID-19 pandemic. Exploratory forecasting was separated from the primary inference because rolling validation showed limited predictive performance.

## Methods

### Study design and data sources

We performed a serial cross-sectional study of US mortality using the CDC WONDER Multiple Cause of Death databases. For 1999-2020, we used the bridged-race multiple-cause files (D77); for 2021-2024, we used the single-race multiple-cause files (D157).[7,8] Both derive from National Vital Statistics System death-certificate files. National overlap analyses for 2018-2020 were extracted from both databases before the series was spliced. C10 and C11 national death counts, populations, crude rates, and AAMRs were identical across databases in each overlap year; the only numerical discrepancy in the audited C10 fields was an age-adjusted rate standard error differing by 0.001 in 2019. Because the data are publicly available, deidentified, and aggregate, the analysis did not involve interaction with human participants or access to identifiable private information. The study is reported in accordance with the STROBE reporting guideline.[13]

### Encephalopathy phenotypes

The primary phenotype was C10, a time-stable multiple-cause definition comprising B22.0, E51.2, G92, G93.1, G93.4, I67.3, I67.4, P57.0, P57.8, and P57.9. C11 added P91.6 and was analyzed from 2006 onward. The narrow sensitivity phenotype was G93.4 alone. B14 added A81.0, A81.1, A81.2, and G93.7 to C10; B15 added P91.6 to B14. Primary analyses identified deaths in which at least one phenotype code appeared among the multiple causes. Separate analyses restricted the phenotype to the underlying cause. Supplementary Table 1 provides the complete definitions and rationale.

### Mortality measures and stratification

We extracted annual deaths, population denominators, crude mortality rates, and directly age-adjusted mortality rates per 100,000 standardized to the 2000 US standard population.[14] We retained CDC-provided confidence intervals and standard errors where available.[15] Age-group analyses used age-specific crude rates rather than age-adjusted rates. Contemporary race-ethnicity analyses used the D157 single-race six-category classification and Hispanic origin; long-term bridged-race analyses were retained separately. We evaluated sex, 10-year and 5-year age groups, race and ethnicity, race by sex, age by race, age by race by sex, state, Census region and division, and urban-rural classification where denominators were supported.

### Data quality, suppression and non-rate outcomes

CDC WONDER suppresses death counts below 10 and identifies statistically unreliable rates according to its published rules.[7,8] Suppressed counts were never imputed as zero. Unreliable rates were preserved as status flags and excluded from rate-based modeling. Suppressed and zero rows were not displayed in the high-cardinality detailed underlying-cause query; CDC notes that hidden rows remain incorporated in applicable totals. Multiple-cause code rows can overlap within a death certificate and were therefore not summed to estimate unique deaths. Place of death, autopsy, weekday, month, and education were treated descriptively when population denominators or rates were not applicable.

### Temporal trend analysis

For the primary national C10 series, we fit continuous piecewise linear regressions to the natural logarithm of annual AAMR using inverse-variance weights derived from the standard error on the log scale. Candidate models contained zero to four internal breakpoints, required a minimum three-year segment, and were selected by Bayesian information criterion. For each segment, we transformed the slope beta to APC = 100 x [exp(beta) − 1] and derived Wald 95% confidence intervals. The average annual percent change was computed as the duration-weighted mean log slope across segments. This approach is conceptually related to joinpoint regression but does not reproduce the NCI permutation-test algorithm; we therefore describe it as weighted segmented log-linear regression rather than official Joinpoint.[16] NCI Joinpoint-ready input files are provided separately.

### Disparity analyses

For sex and contemporary race-ethnicity comparisons, we calculated absolute rate differences and rate ratios. Standard errors for differences were obtained from the square root of the summed rate variances. Rate-ratio confidence intervals were computed on the log scale using the delta method. Non-Hispanic White people were the reference group for contemporary race-ethnicity contrasts, and females were the reference for sex contrasts. Intersectional strata were summarized using observed deaths and crude rates and were not used to infer individual-level effects.

### Underlying-cause and subtype decomposition

Detailed underlying causes among C10-associated deaths were grouped into clinically interpretable ICD-10 chapter-based categories, including circulatory/cerebrovascular, nervous system, respiratory, neoplasm, endocrine/metabolic/nutritional, digestive/hepatic, external/toxic, infectious, renal/genitourinary, perinatal, and COVID-19 categories. Each category was expressed as a proportion of annual C10-associated deaths. Because detailed rows below the suppression threshold were hidden, an unclassified remainder captured the difference between national totals and displayed categories. Individual C10 multiple-cause codes were analyzed separately; exploratory long-term log-linear slopes were adjusted across codes using the Benjamini-Hochberg false-discovery-rate procedure.

### COVID-19 and secondary analyses

For 2020-2024, we quantified deaths meeting C11 in which U07.1 was the underlying cause and expressed these as a fraction of annual C11-associated deaths. Supplementary analyses evaluated infant-age strata, perinatal etiologies, urban-rural patterns, place of death, autopsy, weekday, month, and education. Some historical extracts contained additional sex or age dimensions relative to their baseline query definitions; the analytic pipeline preserved the extracted structure and labeled those outputs accordingly in the supplementary files.

### Exploratory forecasting

Forecasting was prespecified as exploratory and excluded from the primary inferential claims. We compared naive, log-linear drift, damped Holt, and ARIMA models using rolling-origin validation and summarized performance with mean absolute error, root mean squared error, and mean absolute scaled error. Although ARIMA had the lowest mean absolute error, all models had MASE greater than one and the naive model had lower RMSE than ARIMA. We therefore present 2025-2030 projections only in Supplementary Information as conditional scenarios. A separate age-specific scenario combined projected age-specific rates with US Census population projections to decompose changes into population-composition and rate effects.

## Results

### National mortality followed four distinct temporal phases

Across 1999-2024, the primary C10 definition identified 953,846 encephalopathy-associated deaths. Annual deaths increased from 25,055 in 1999 to 49,334 in 2024, with the highest annual count in 2021 (56,980). The age-adjusted mortality rate (AAMR) increased from 9.143 per 100,000 (95% CI, 9.029-9.256) in 1999 to 11.738 (11.633-11.845) in 2024, a 28.4% net increase, and peaked at 14.391 (14.271-14.513) in 2021 (Fig. 1a,b; Table 1).

**Figure 1.**
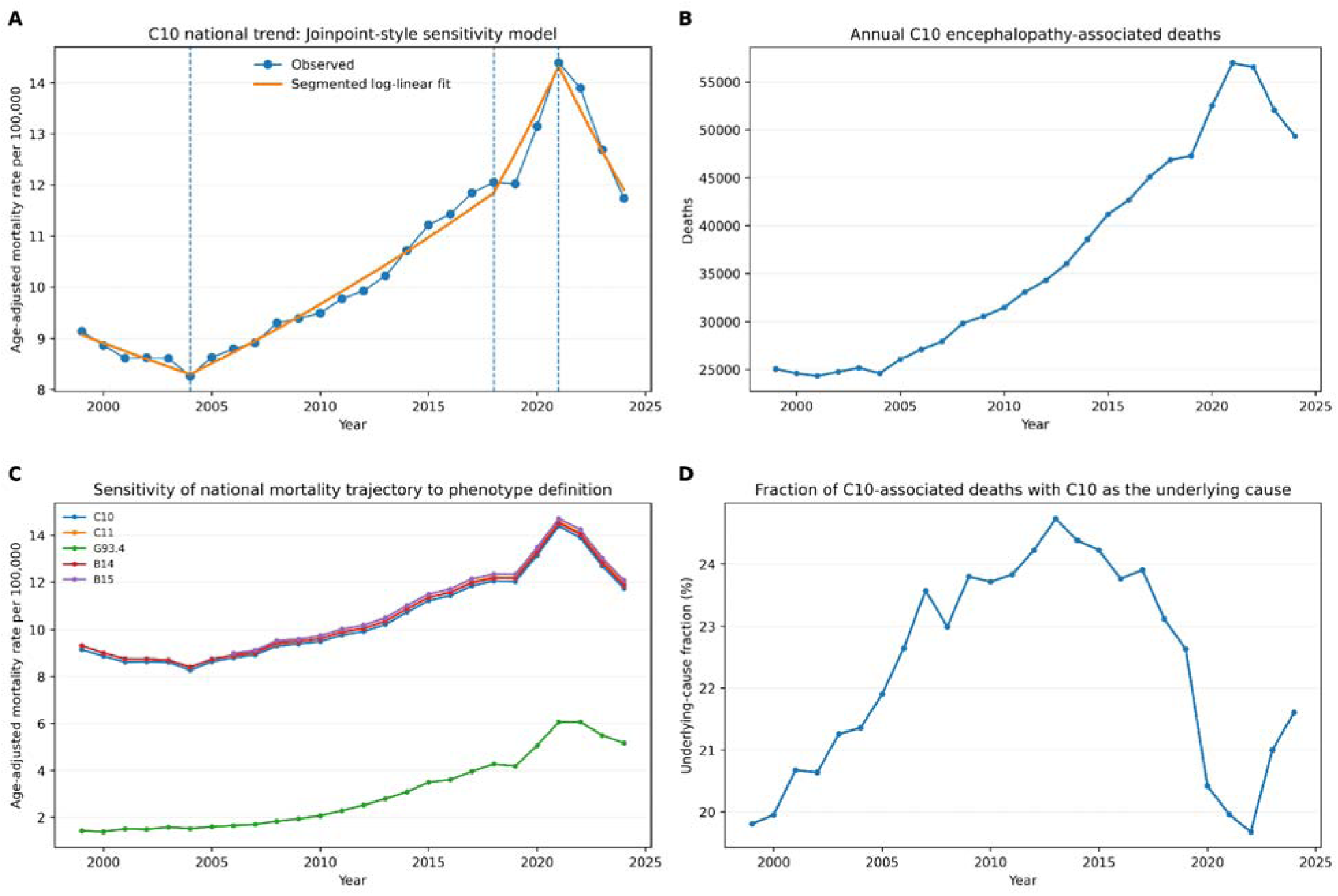
National trajectory and phenotype robustness. a, Observed C10 national AAMRs and the weighted segmented log-linear fit; vertical dashed lines mark data-driven breakpoints. b, Annual C10-associated deaths. c, National AAMR trajectories across nested phenotypes (C10, C11, G93.4, B14 and B15). d, Percentage of C10-associated deaths in which a C10 code was also the underlying cause. Rates are per 100,000 population.

**Table 1.** National C10 benchmark years.

| Year | Deaths | Population | Crude rate | AAMR (95% CI) |
| --- | --- | --- | --- | --- |
| 1999 | 25,055 | 279,040,168 | 8.979 | 9.143 (9.029-9.256) |
| 2006 | 27,091 | 298,379,912 | 9.079 | 8.796 (8.691-8.901) |
| 2019 | 47,280 | 328,239,523 | 14.404 | 12.021 (11.910-12.131) |
| 2020 | 52,494 | 329,484,123 | 15.932 | 13.147 (13.032-13.262) |
| 2021 | 56,980 | 331,893,745 | 17.168 | 14.391 (14.271-14.513) |
| 2024 | 49,334 | 340,110,988 | 14.505 | 11.738 (11.633-11.845) |

AAMR, age-adjusted mortality rate per 100,000; crude rates are per 100,000.

Weighted segmented log-linear regression selected breakpoints at 2004, 2018, and 2021. The AAMR declined during 1999-2004 (annual percent change [APC], −1.76%; 95% CI, −2.64% to −0.88%), increased during 2004-2018 (APC, 2.57%; 2.33%-2.81%), accelerated during 2018-2021 (APC, 6.54%; 5.37%-7.72%), and declined during 2021-2024 (APC, −5.98%; −7.34% to −4.61%). The duration-weighted average annual percent change across the full interval was 1.09% (95% CI, 0.91%-1.28%) (Fig. 1a; Table 3).

### The national trajectory was robust to nested phenotype definitions but not compositionally static

C10, C11, and the broader B14/B15 phenotypes produced closely aligned national trajectories, including the 2021 peak and subsequent decline (Fig. 1c). The narrow G93.4 phenotype increased from an AAMR of 1.419 per 100,000 in 1999 to 5.168 in 2024. The comprehensive C11 phenotype, available from 2006 because P91.6 entered the classification in that period, had an AAMR of 11.944 in 2024; B15 was 12.077.

Most C10-associated deaths did not list a C10 code as the underlying cause. The underlying-cause fraction was 19.8% in 1999, rose to approximately one quarter in the early 2010s, and was 21.6% in 2024 (Fig. 1d). An underlying-cause-only definition would therefore have captured only a minority of deaths in which the prespecified encephalopathy phenotype was documented.

### Mortality burden was concentrated at older ages and showed persistent sex and race-ethnicity differences

Age-specific mortality increased steeply with age. In 2024, the C10 crude mortality rate was 121.536 per 100,000 among people aged 85 years or older, 59.902 at ages 75-84 years, 33.982 at ages 65-74 years, and 20.873 at ages 55-64 years (Fig. 2b). Rates were substantially lower at younger ages, although infants formed a distinct small high-risk group relative to older children.

**Figure 2.**
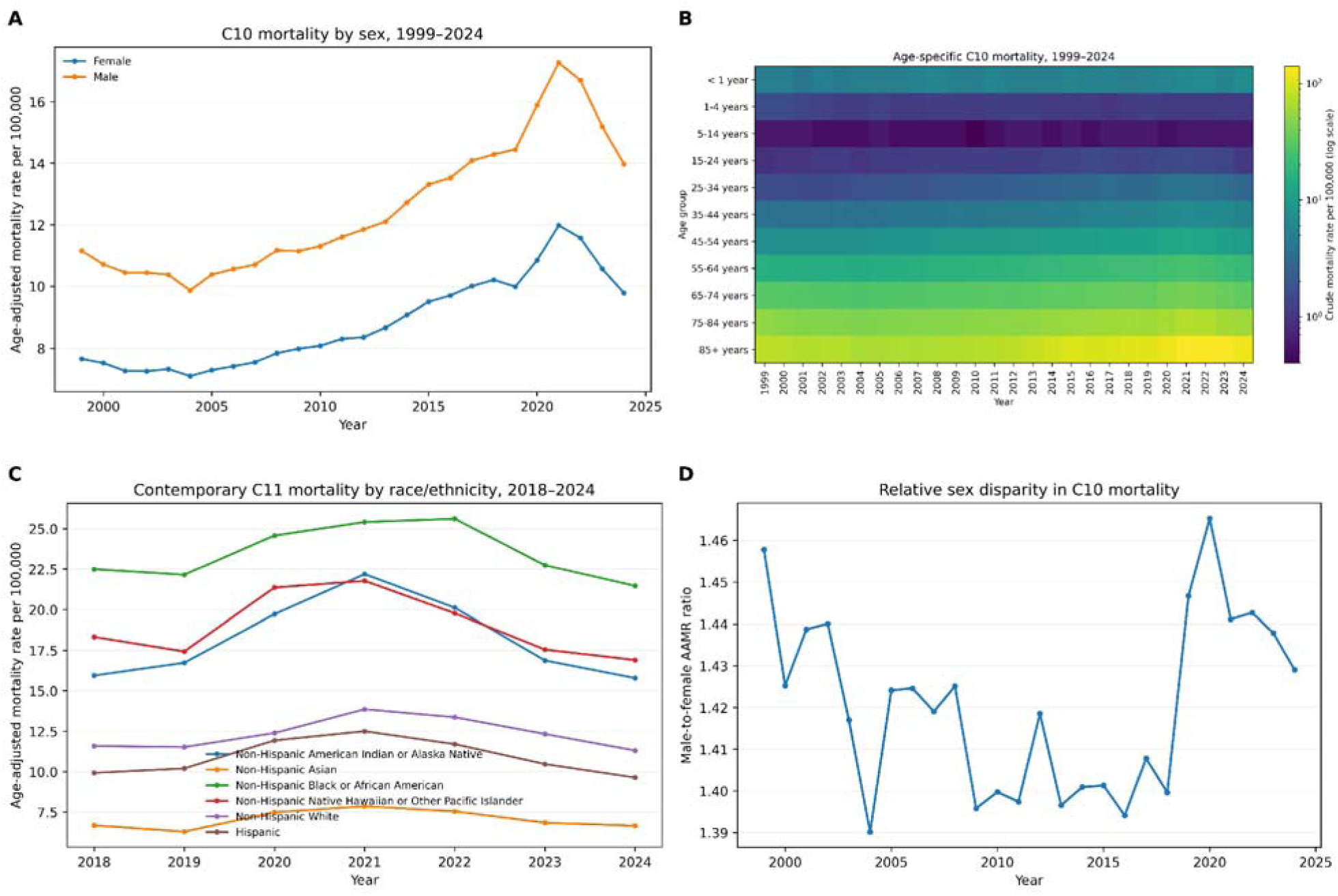
Demographic patterning of encephalopathy-associated mortality. a, C10 AAMR by sex, 1999-2024. b, Heat map of age-specific C10 crude mortality rates. c, C11 AAMR by contemporary race and ethnicity, 2018-2024. d, Male-to-female C10 AAMR ratio over time with 95% confidence intervals. Race-ethnicity categories use the single-race D157 classification for contemporary analyses.

Sex differences persisted across the study period (Fig. 2a,d). In 1999, the male AAMR was 11.149 per 100,000 compared with 7.648 in females (rate ratio [RR], 1.46; 95% CI, 1.42-1.49). In 2024, the corresponding rates were 13.973 and 9.778 (RR, 1.43; 1.40-1.46), yielding an absolute rate difference of 4.195 per 100,000. The relative difference remained broadly stable while the absolute gap widened during the pandemic-era peak.

Contemporary single-race analyses using C11 showed substantial differences across race and ethnicity (Fig. 2c; Table 2). In 2024, the AAMR among non-Hispanic Black people was 21.466 per 100,000 compared with 11.308 among non-Hispanic White people (RR, 1.90; 95% CI, 1.85-1.94). Rates were also higher than the White reference among non-Hispanic Native Hawaiian or Other Pacific Islander people (16.898; RR, 1.49) and non-Hispanic American Indian or Alaska Native people (15.784; RR, 1.40), whereas rates were lower among Hispanic people (9.638; RR, 0.85) and non-Hispanic Asian people (6.650; RR, 0.59). Intersectional analyses showed substantial age-dependent variation within race-ethnicity and sex strata (Fig. 3).

**Figure 3.**
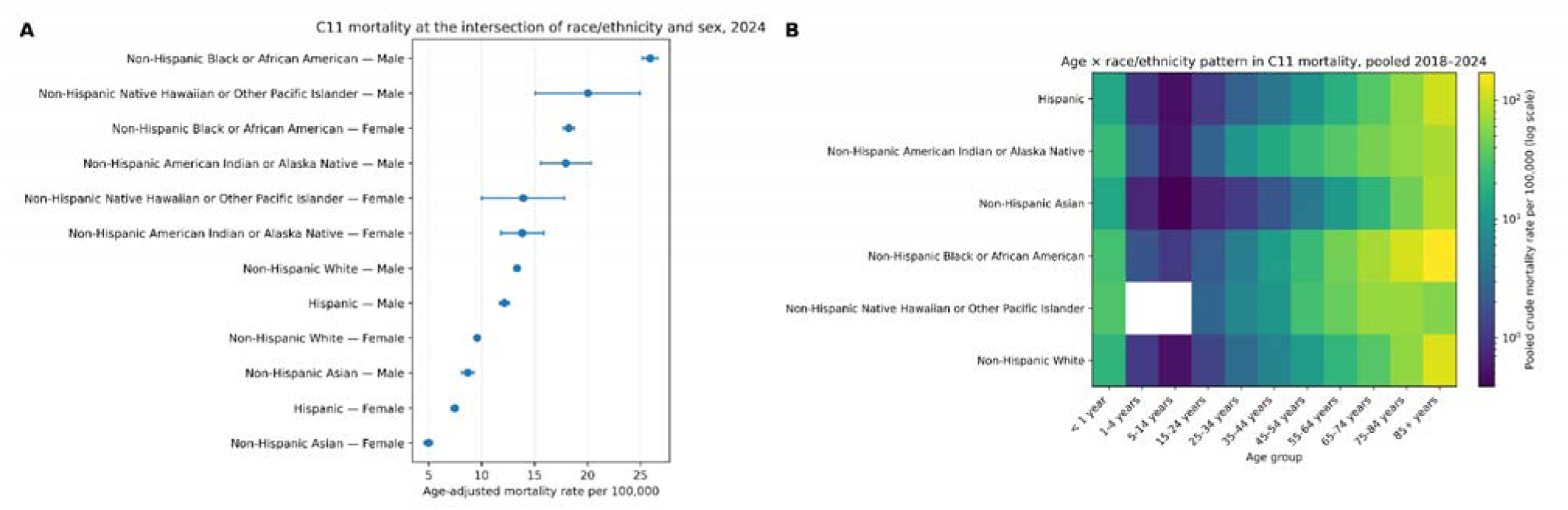
Intersectional demographic heterogeneity. a, C11 mortality by race-ethnicity and sex in 2024. b, Pooled 2018-2024 age-by-race-ethnicity patterning. These analyses are descriptive and do not identify causal mechanisms underlying group differences.

**Table 2.** Demographic AAMR snapshot in 2024.

| Dimension | Group | Phenotype | Deaths | AAMR |
| --- | --- | --- | --- | --- |
| Sex | Female | C10 | 22,266 | 9.778 |
| Sex | Male | C10 | 27,068 | 13.973 |
| Race/ethnicity | Non-Hispanic AIAN | C11 | 420 | 15.784 |
| Race/ethnicity | Non-Hispanic Asian | C11 | 1,581 | 6.650 |
| Race/ethnicity | Non-Hispanic Black | C11 | 9,584 | 21.466 |
| Race/ethnicity | Non-Hispanic NHOPI | C11 | 115 | 16.898 |
| Race/ethnicity | Non-Hispanic White | C11 | 32,534 | 11.308 |
| Race/ethnicity | Hispanic | C11 | 5,105 | 9.638 |

AIAN, American Indian or Alaska Native; NHOPI, Native Hawaiian or Other Pacific Islander.
Contemporary race-ethnicity estimates use the 2018-2024 single-race database.

**Table 3.** Weighted segmented log-linear C10 trend.

| Period | APC, % | 95% CI | P value |
| --- | --- | --- | --- |
| 1999-2004 | -1.76 | -2.64 to -0.88 | <0.001 |
| 2004-2018 | 2.57 | 2.33 to 2.81 | <0.001 |
| 2018-2021 | 6.54 | 5.37 to 7.72 | <0.001 |
| 2021-2024 | -5.98 | -7.34 to -4.61 | <0.001 |

APC, annual percent change. Breakpoints were selected by Bayesian information criterion in the prespecified segmented-regression sensitivity framework; this is not the official NCI Joinpoint permutation-test algorithm.

### Geographic heterogeneity persisted throughout the study period

Regional trajectories diverged over time (Fig. 4c). In 2024, C10 AAMRs were 13.144 per 100,000 in the Midwest, 13.120 in the South, 10.415 in the West, and 8.730 in the Northeast. State-level estimates showed broader dispersion than regional averages (Fig. 4a). The coefficient of variation across state AAMRs was 0.328 in 1999, increased to 0.359 in 2021, and was 0.303 in 2024; the 90th-to-10th percentile ratio was 1.85, 2.50, and 2.19 at those time points, respectively (Fig. 4b).

**Figure 4.**
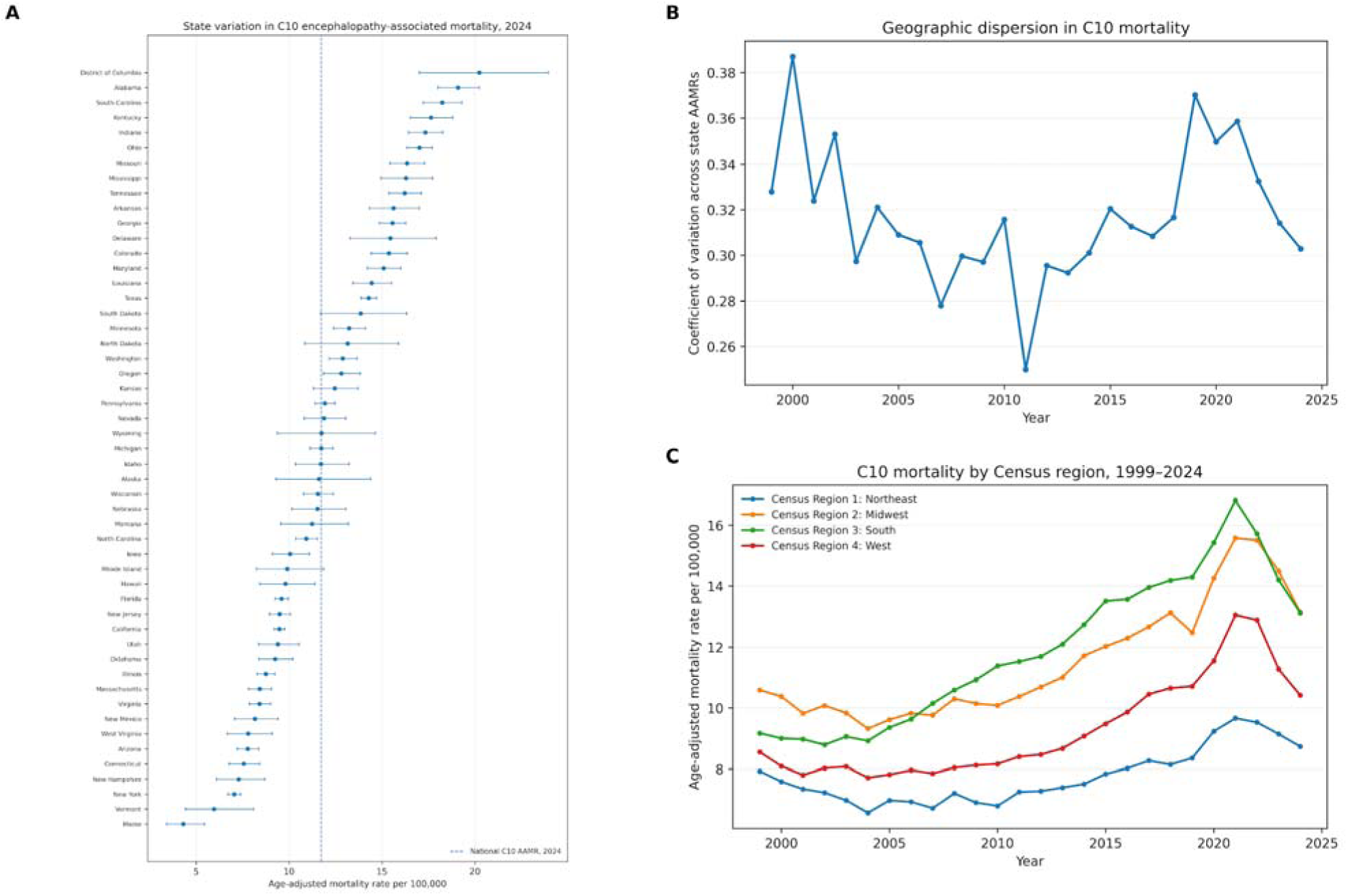
Geographic heterogeneity in C10 encephalopathy-associated mortality. a, State-specific 2024 AAMRs with 95% confidence intervals; the dashed vertical line marks the 2024 national C10 AAMR (11.738 per 100,000). b, Coefficient of variation across state AAMRs over time. c, AAMRs by US Census region, 1999-2024.

### The clinical and underlying-cause composition changed over time

The national trend was not accompanied by uniform increases in every component code (Fig. 5a). Anoxic brain damage (G93.1) remained the largest individual C10 component, with an AAMR of 7.556 in 1999 and 6.282 in 2024. By contrast, unspecified encephalopathy (G93.4) increased from 1.419 to 5.168; Wernicke encephalopathy (E51.2) increased from 0.013 to 0.132; and toxic encephalopathy (G92) increased from 0.037 to 0.132. Exploratory code-specific log-linear trends, corrected using the Benjamini-Hochberg procedure, supported increases for G93.4, E51.2, G92, and I67.4, while the G93.1 long-term slope was comparatively small. Because multiple-cause codes can co-occur on the same certificate, code-specific rows were not summed to estimate unique deaths.

**Figure 5.**
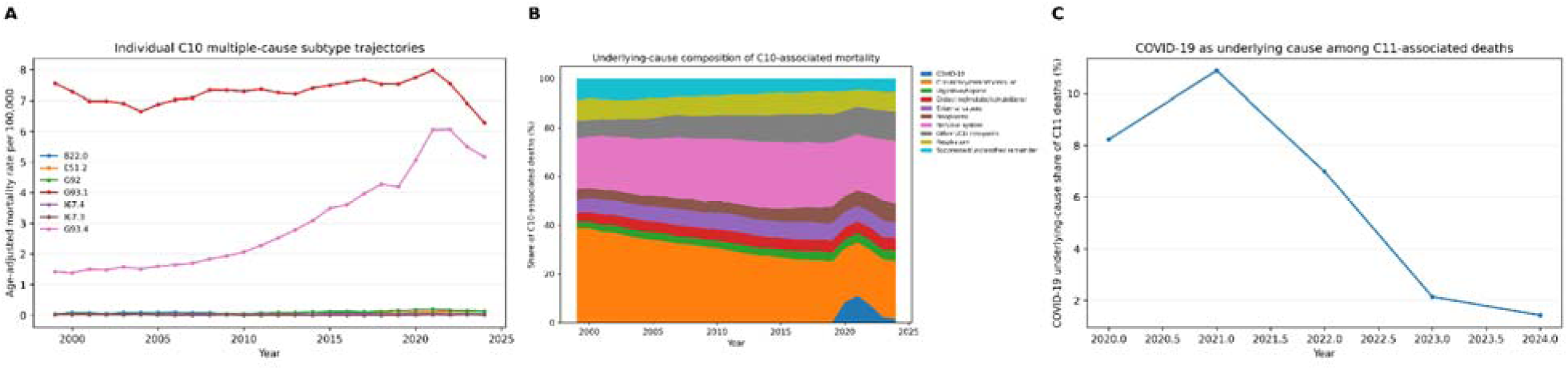
Changing diagnostic and underlying-cause composition. a, AAMR trajectories for individual C10 multiple-cause codes. b, Distribution of underlying-cause categories among C10-associated deaths over time. c, Percentage of C11-associated deaths with COVID-19 (U07.1) as the underlying cause, 2020-2024. Multiple-cause code rows can overlap within a death and are not additive.

The underlying diseases accompanying C10-associated deaths also shifted (Fig. 5b). Circulatory or cerebrovascular causes accounted for 38.7% of C10-associated deaths in 1999 and 23.9% in 2024, while nervous-system causes accounted for 20.9% and 25.8%, respectively. COVID-19 was the underlying cause in 8.24% of C11-associated deaths in 2020 and 10.98% in 2021; the share fell to 6.98% in 2022, 2.14% in 2023, and 1.42% in 2024 (Fig. 5c).

Secondary analyses of urban-rural patterns, infant-age strata, place of death, autopsy status, month, weekday, and education are presented in Supplementary Information. Recent single-race urban-rural denominators were not available for all years, so those outputs were kept descriptive where rates were unsupported. Exploratory 2025-2030 projections were also retained in the supplement: ARIMA minimized mean absolute error, but all candidate models had mean absolute scaled error greater than one and the naive model had a lower RMSE, indicating limited forecasting skill.

## Discussion

Across 1999-2024, encephalopathy-associated mortality followed a nonlinear national trajectory, with sustained pre-pandemic growth, a 2021 peak, and a partial decline by 2024. The direction of change was consistent across nested syndrome definitions despite a marked shift toward unspecified encephalopathy. Mortality remained concentrated at older ages and differed by sex, race-ethnicity, and geography, while the pandemic period temporarily altered the distribution of underlying causes.

The distinction between a stable syndrome-level trajectory and changing code composition is important. Encephalopathy encompasses mechanistically diverse conditions, and no single ICD-10 code captures the construct.[1–6] The rise in G93.4 could reflect changes in disease occurrence, recognition, documentation, coding practice, migration from more specific labels, or a combination of these factors. The relative long-term stability of anoxic brain damage argues against interpreting the total increase as a uniform expansion of every encephalopathy mechanism. The nested phenotype analyses therefore serve as a measurement sensitivity analysis: the broad national trend persisted, but its diagnostic composition changed.

The multiple-cause framework materially changes surveillance estimates. Only about one fifth of C10- associated deaths listed a C10 code as the underlying cause in 2024. Underlying-cause statistics identify the disease or injury initiating the chain leading to death, whereas multiple-cause data also retain conditions recorded as contributing.[7,8] For encephalopathy, which often occurs during severe systemic or neurologic illness, this broader documentation is epidemiologically informative. These estimates describe death certificates on which the phenotype was recorded and do not establish that encephalopathy caused every associated death.

The age gradient is clinically plausible because older adults carry greater burdens of cerebrovascular disease, cardiac arrest, renal or hepatic dysfunction, infection, medication exposure, and other precipitants of global cerebral dysfunction.[1,2] The persistence of a roughly 1.4- to 1.5-fold male-to-female AAMR ratio suggests that the sex difference was not confined to the pandemic peak. Race-ethnicity contrasts require additional context: mortality was highest among non-Hispanic Black people in the contemporary C11 analysis, with elevated rates also observed among American Indian or Alaska Native and Native Hawaiian or Other Pacific Islander groups. The death-certificate data do not include socioeconomic conditions, comorbidity severity, care pathways, or treatment exposure, and race classification can be imperfect, particularly for American Indian or Alaska Native decedents.[9] These patterns therefore identify populations for further etiologic study rather than mechanisms of disparity.

Geographic heterogeneity also persisted across regions and states. The South and Midwest had the highest regional AAMRs in 2024, and state dispersion remained substantial after the post-2021 decline. Similar geographic variation has been reported in other CDC WONDER analyses of neurologic and multiorgan mortality phenotypes.[11,12] State-level vital statistics combine differences in population structure, disease prevalence, health-system practice, and certification. Linking mortality phenotypes to hospital data, comorbidity profiles, social determinants, and certifier-level coding patterns would help distinguish these explanations.

The pandemic-era increase was pronounced but transient. Neurologic dysfunction, impaired consciousness, cerebrovascular events, and encephalopathy were recognized among patients with severe COVID-19 early in the pandemic.[17,18] In our data, COVID-19 was the underlying cause for 8.2% of C11-associated deaths in 2020 and 10.9% in 2021, falling to 1.4% by 2024. This temporal pattern is compatible with both direct and indirect pandemic effects, including infection severity, treatment and vaccination, hospital strain, changes in coding, non-COVID critical illness, and broader population mortality patterns. Death-certificate data cannot separate these pathways.

This study has several strengths: nationwide coverage across the ICD-10 era through 2024; nested phenotype definitions rather than reliance on a single code; explicit separation of underlying and contributing causes; empirical validation of the 2018-2020 transition from bridged-race to single-race WONDER data; analysis of both absolute and relative demographic disparities; and decomposition of syndrome codes and underlying causes. The analytic pipeline also preserved suppression and unreliability flags, avoided imputing suppressed counts as zero, and generated source data and code for reproducibility. Identical C10 and C11 national death counts and rates across the overlap years supported the national splice used for 2021-2024.

Several limitations should be considered. Death certificates are administrative clinical records rather than adjudicated neurologic diagnoses, and cause-of-death certification errors are well documented.[10] The C10 phenotype was designed to balance specificity and temporal stability but is not a validated clinical case definition; broad sensitivity phenotypes may include distinct diseases, whereas narrow definitions may miss relevant syndromes. Changes in diagnostic awareness and coding can mimic epidemiologic trends. P91.6 was analyzed only from 2006 onward, preventing a uniform C11 series across the full period. Suppression reduces detail in small cells, some recent urban-rural population denominators are unavailable in the single-race database, race and ethnicity can be misclassified on death certificates,[9] and state analyses are ecological. The segmented regression is not the NCI Joinpoint permutation-test algorithm.[16] Exploratory forecasts also had limited rolling-origin performance and should be interpreted as conditional scenarios rather than predictions.

In summary, US encephalopathy-associated mortality changed substantially over 1999-2024, but the aggregate increase masks distinct temporal phases and evolving diagnostic composition. Persistent demographic and geographic heterogeneity, together with the large gap between multiple-cause and underlying-cause ascertainment, defines priorities for follow-up. Linking vital statistics to clinical data will be necessary to determine how much of the observed pattern reflects changes in incidence, severity, comorbidity, care delivery, and diagnostic documentation.

## Data availability

All underlying mortality data are publicly available from CDC WONDER.[7,8]

## Data Availability

All data used in this study are publicly available from the Centers for Disease Control and Prevention (CDC) WONDER database at https://wonder.cdc.gov/. The analyses were based on publicly accessible multiple-cause mortality data for the United States.

## Acknowledgements

During the preparation of this article, the authors used ChatGPT (OpenAI) to assist with improving language and readability. The authors reviewed and edited the output as needed and take full responsibility for the content of the published article.

## Author contributions

K.H. conceived the study and designed the analysis. R.M.B. and M.A.S. extracted and curated the data. K.H. performed the statistical analysis and visualization. All authors contributed to the interpretation of the findings. K.H. drafted the manuscript. All authors critically revised the manuscript, approved the final version, and accept responsibility for the integrity of the work.

---

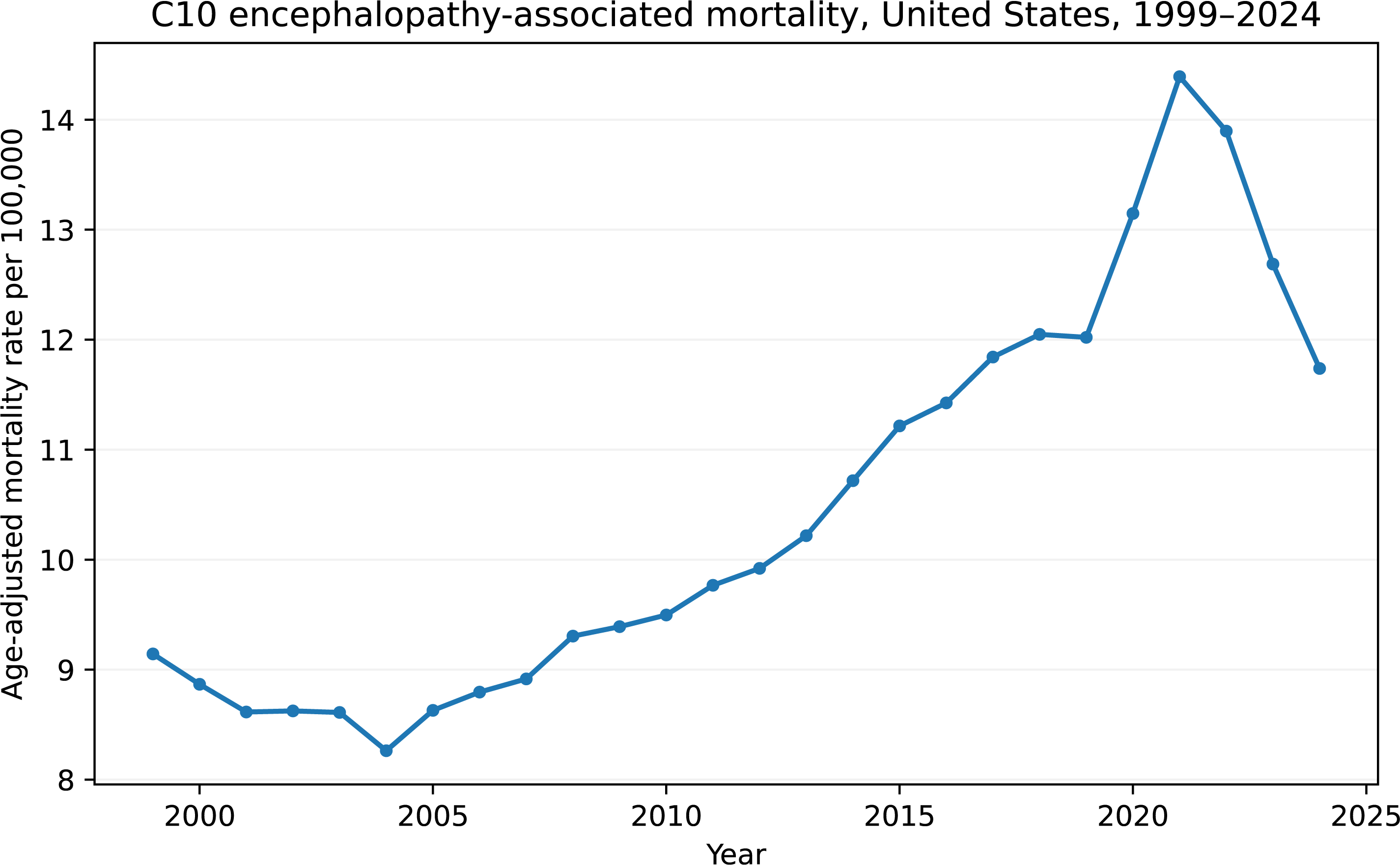

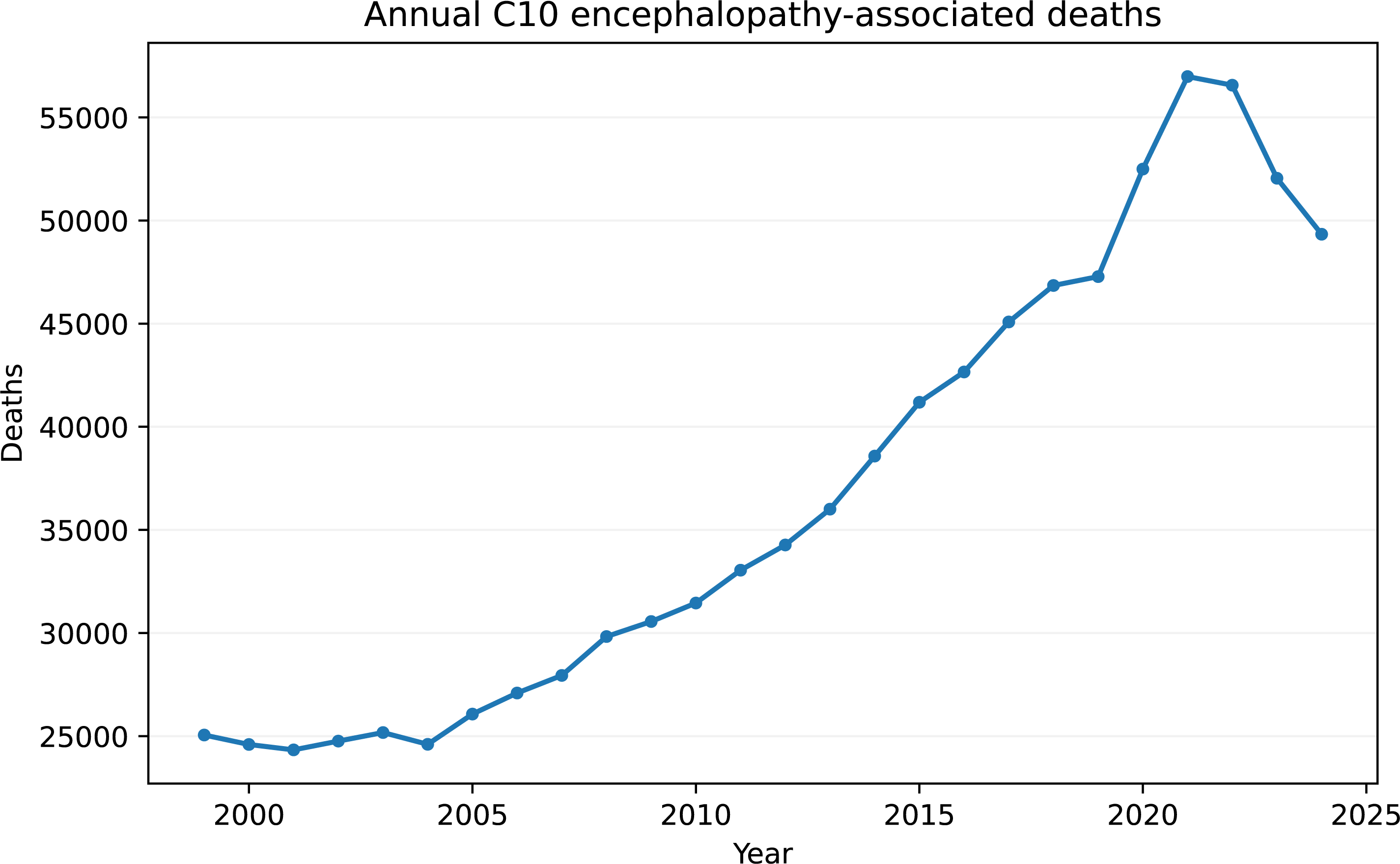

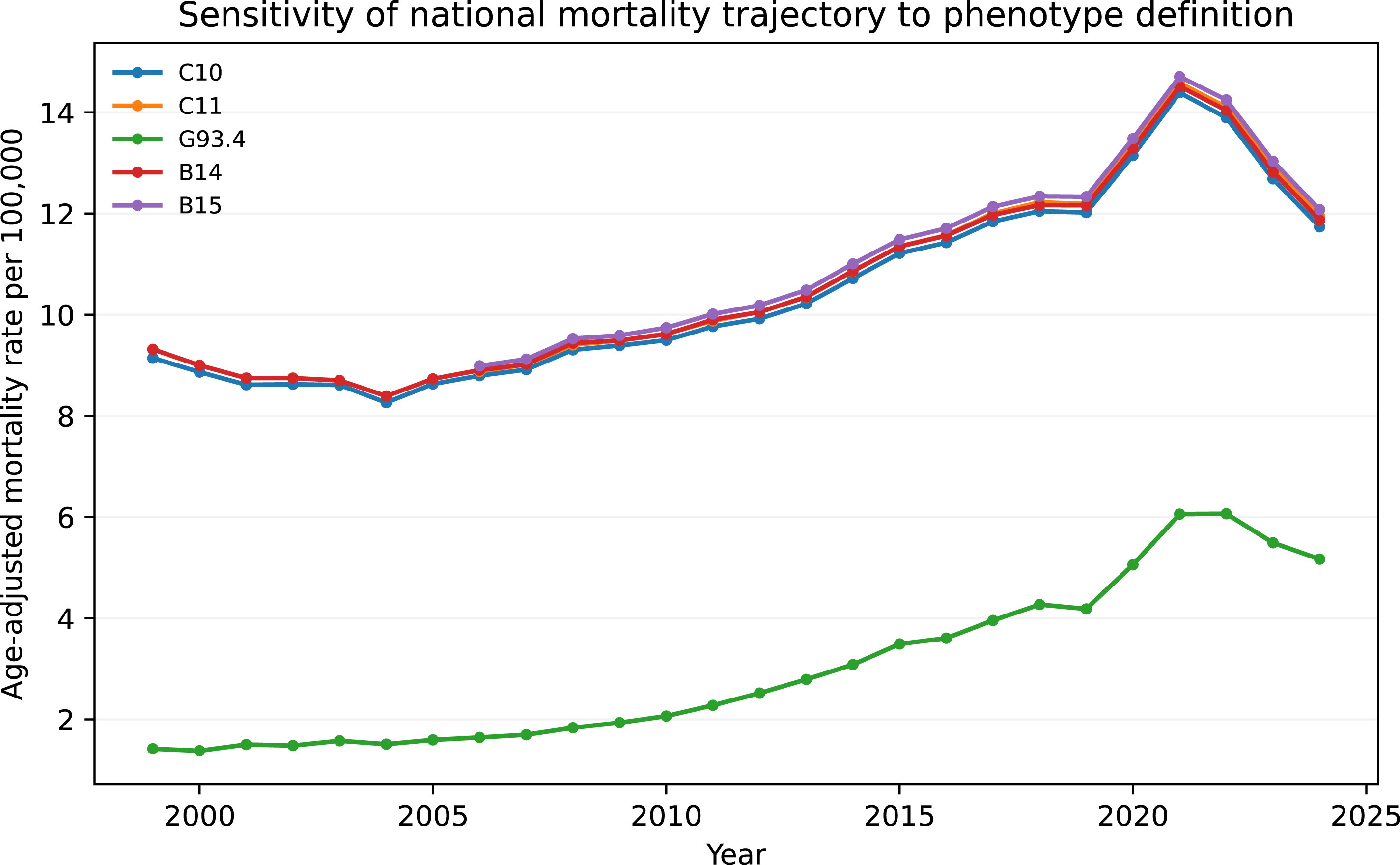

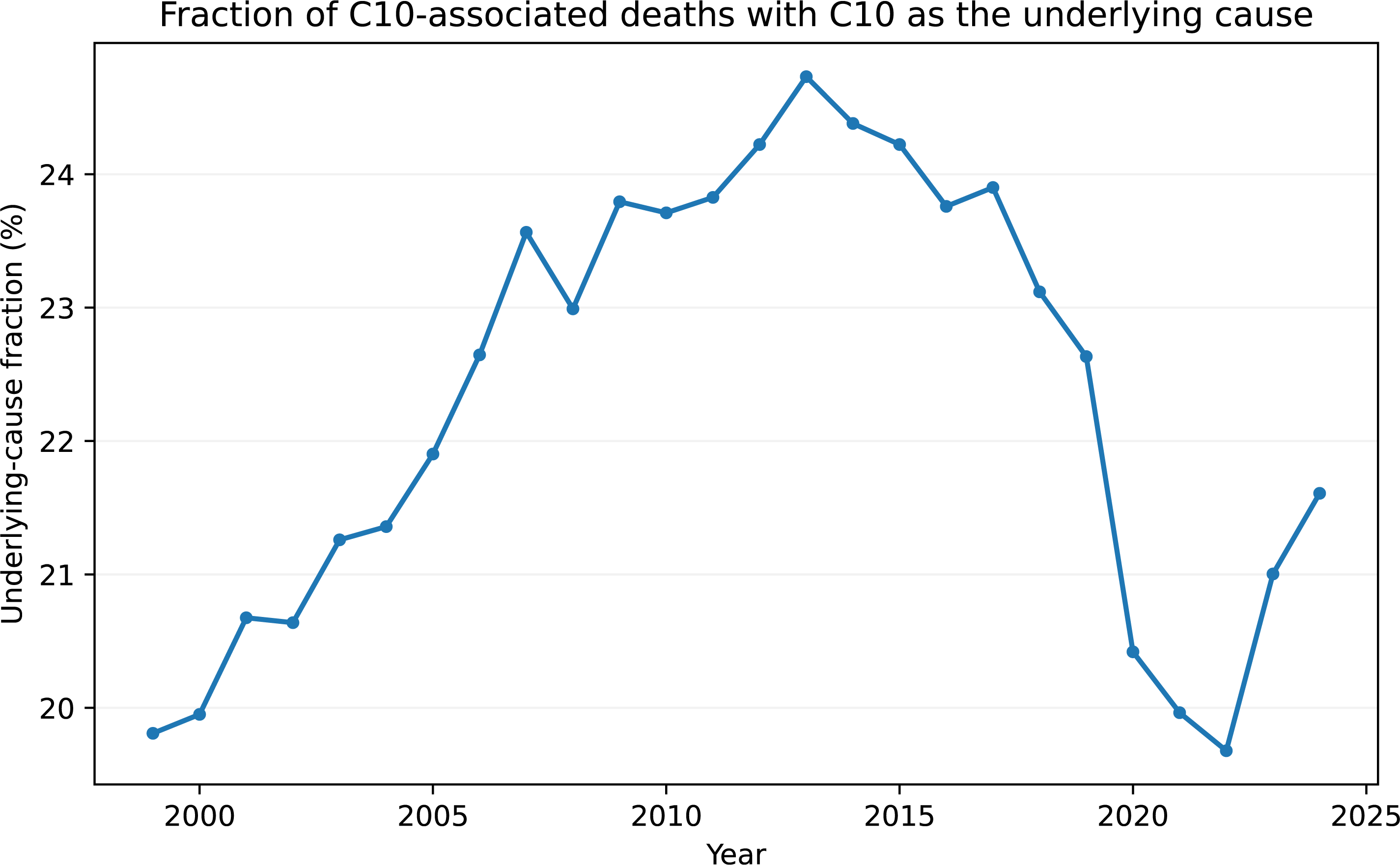

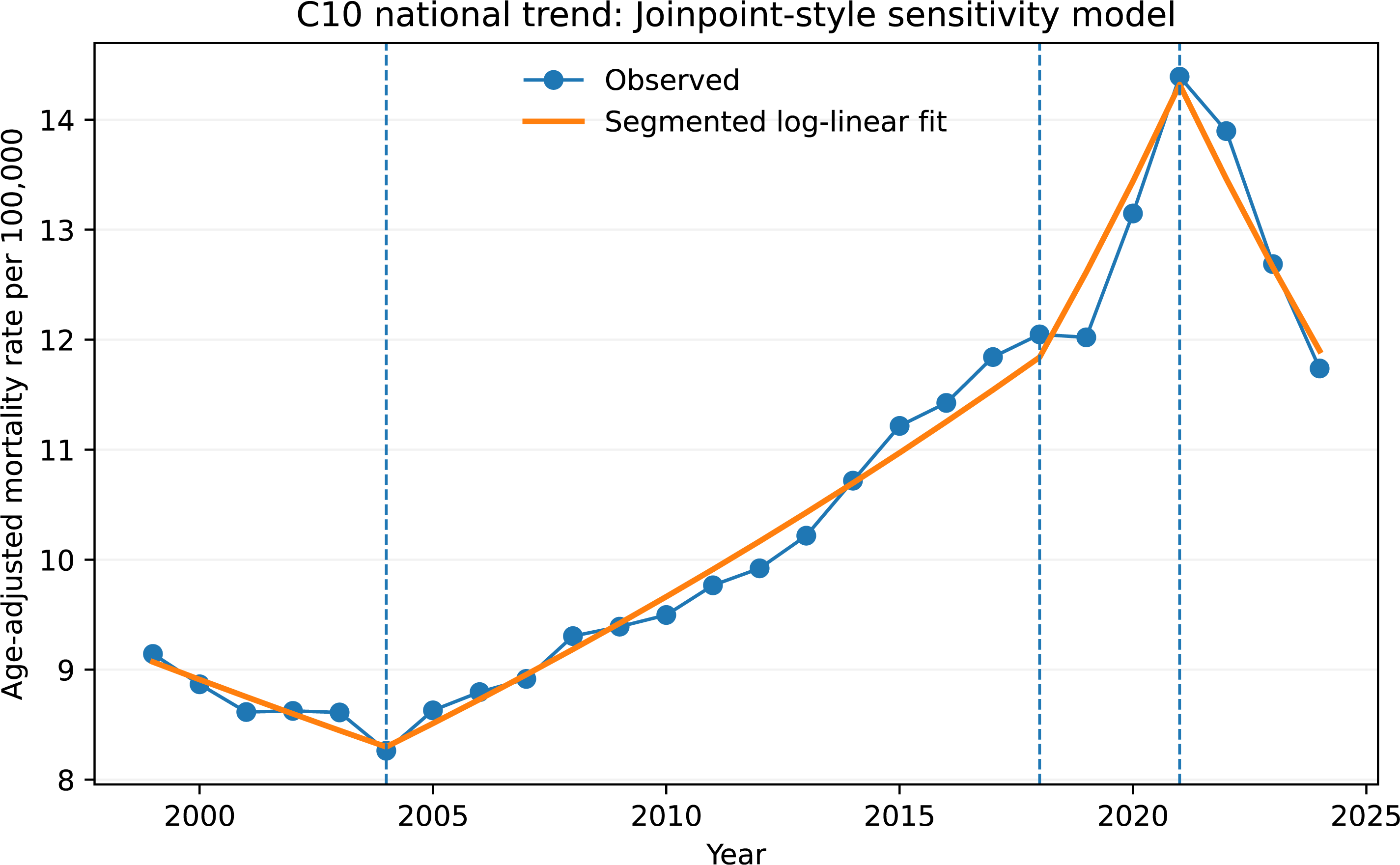

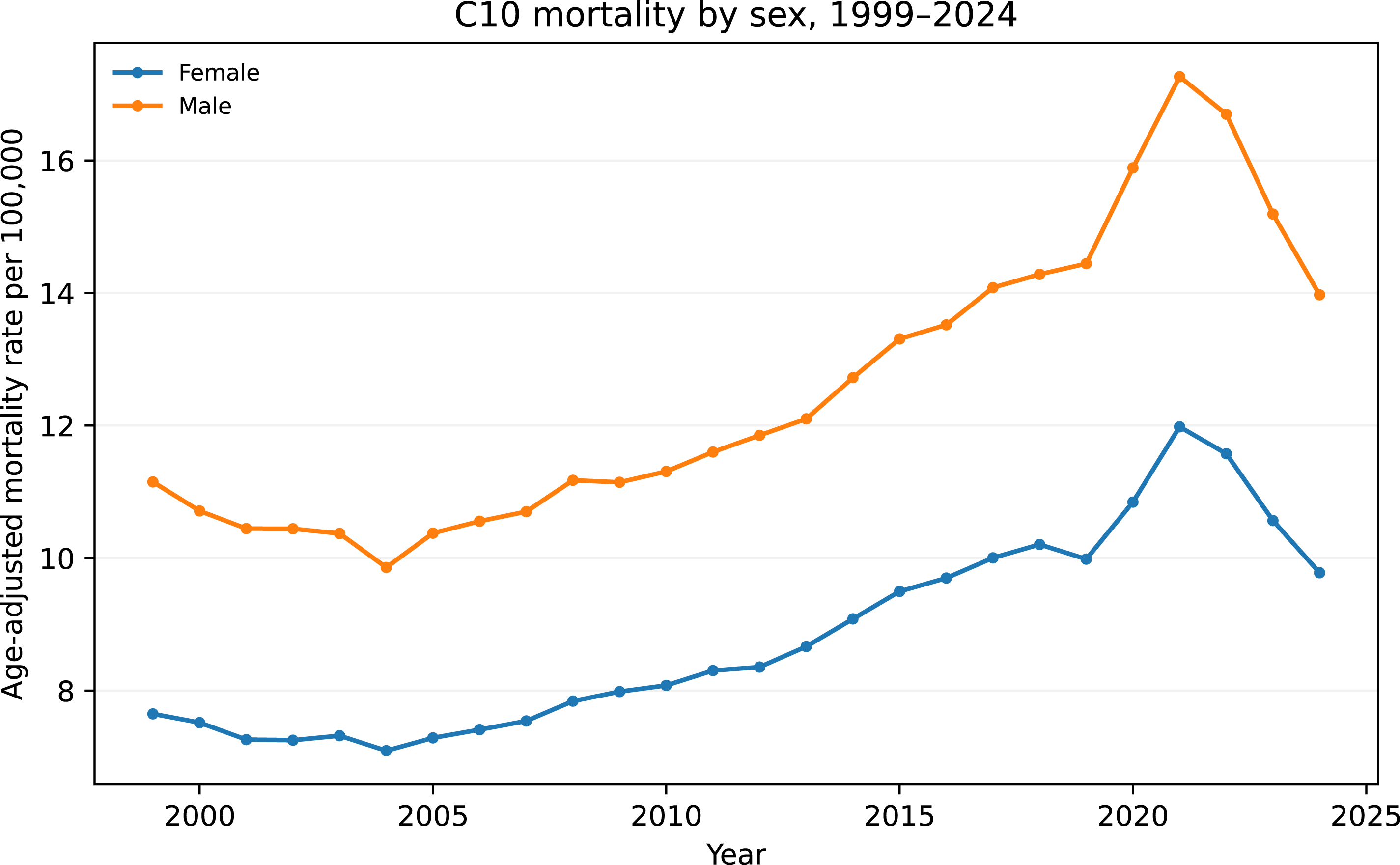

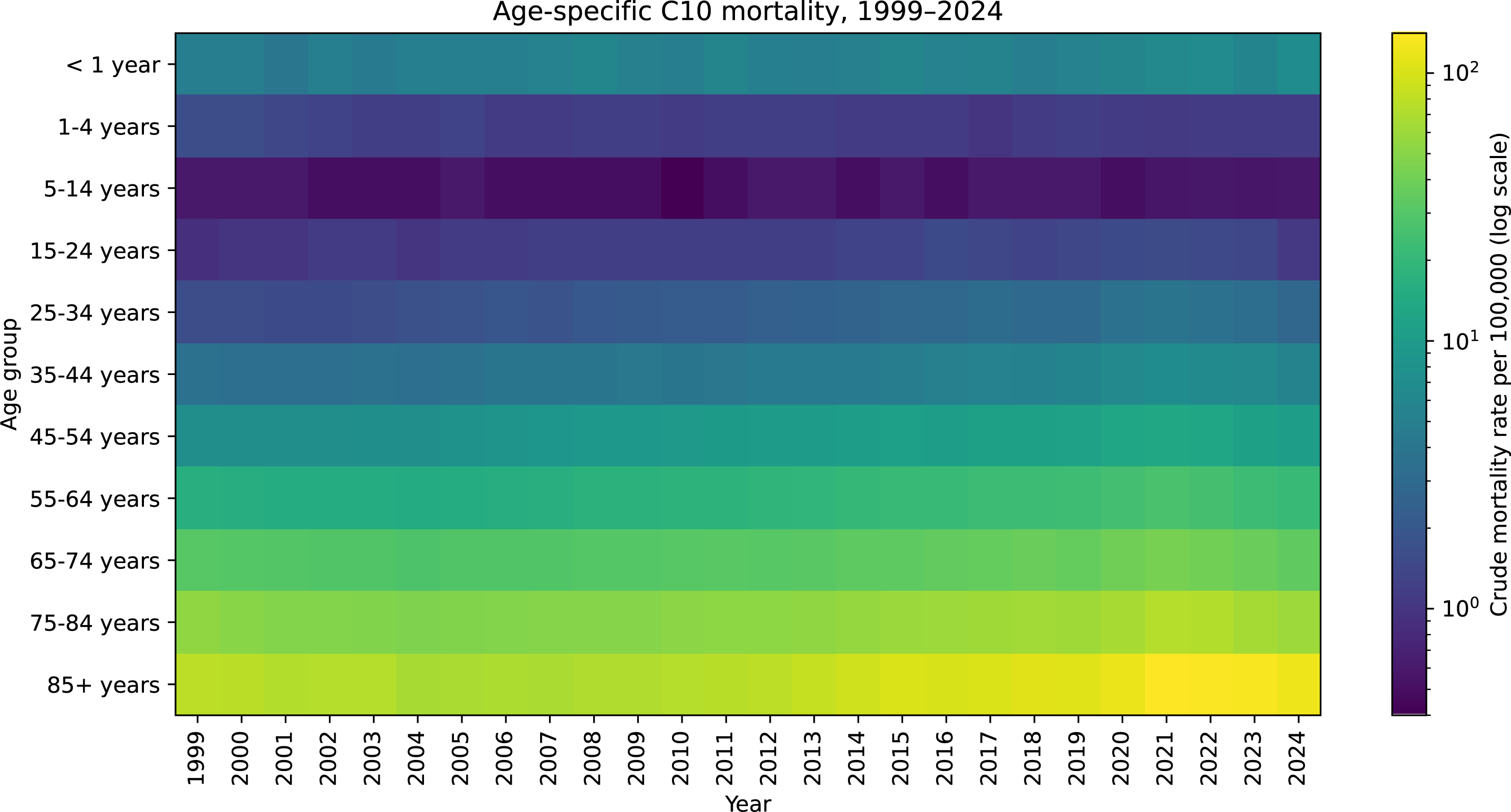

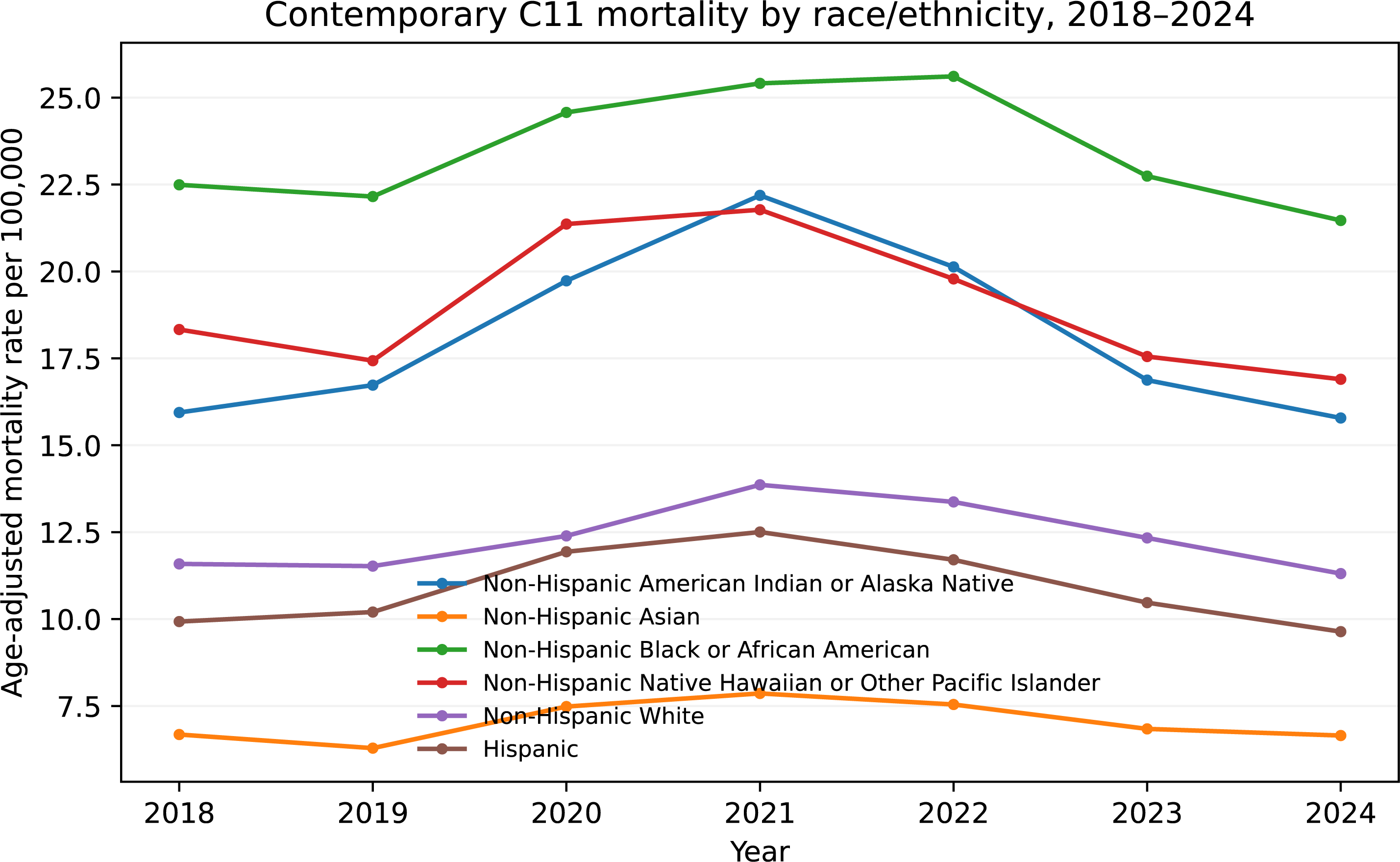

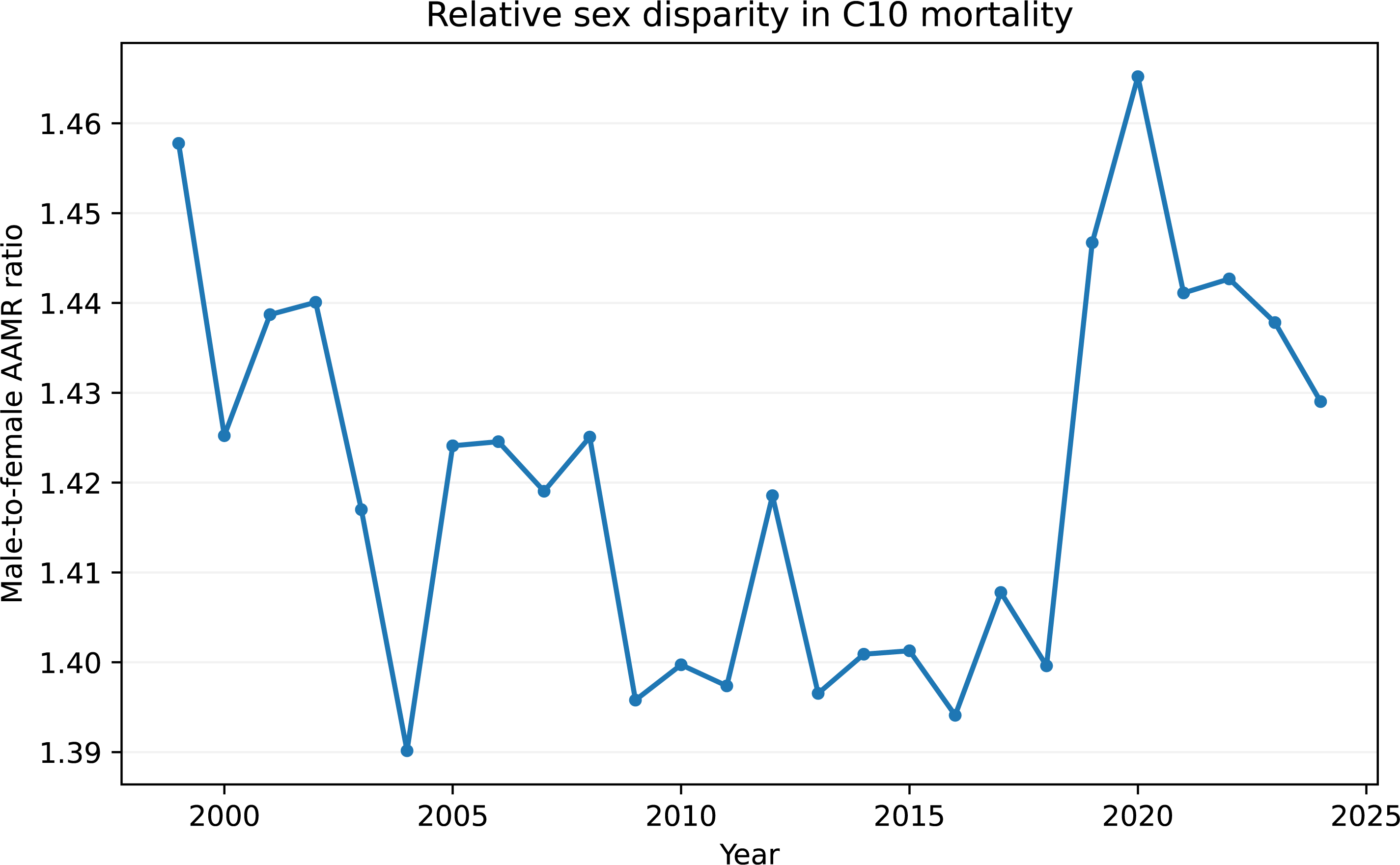

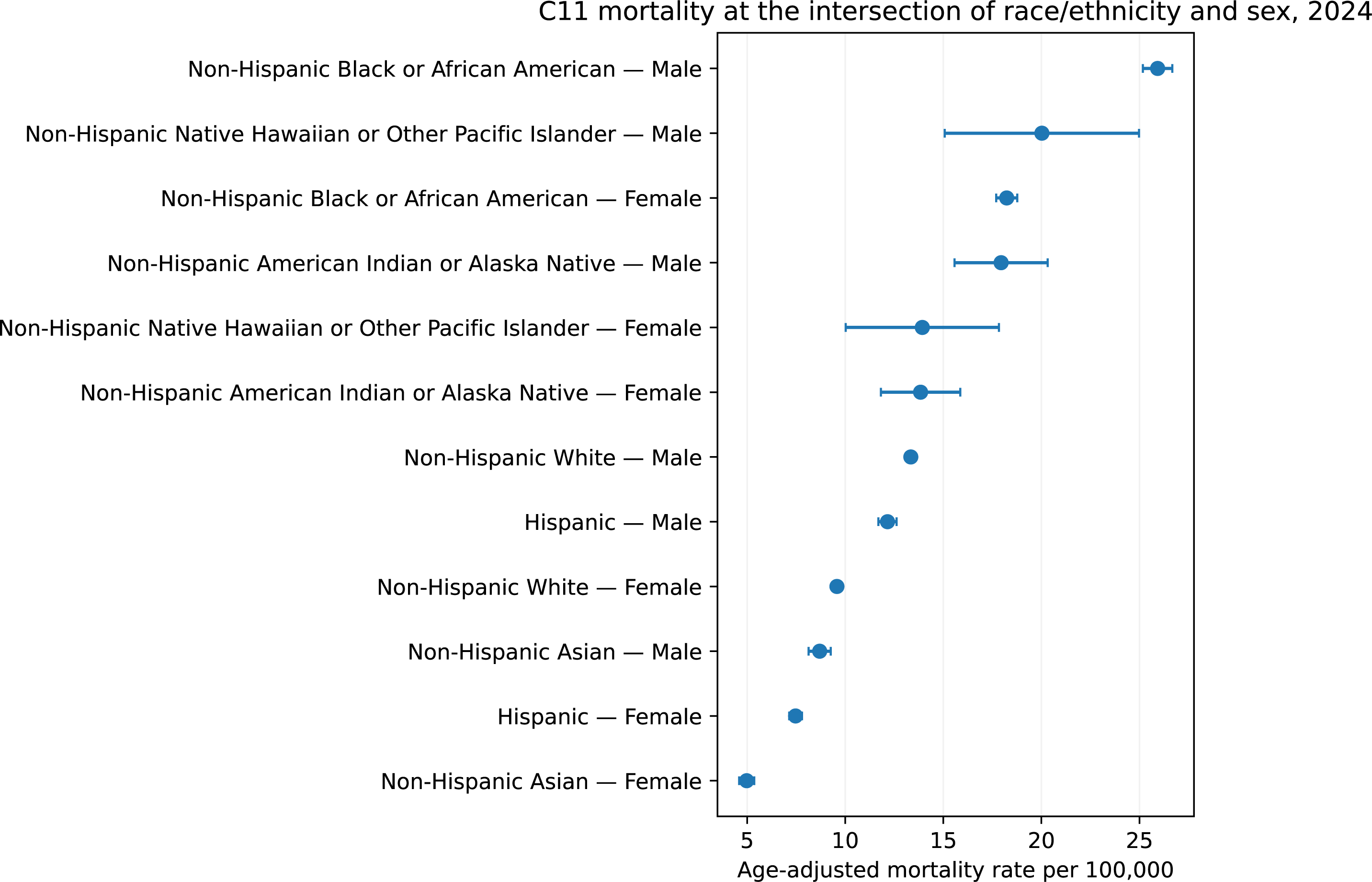

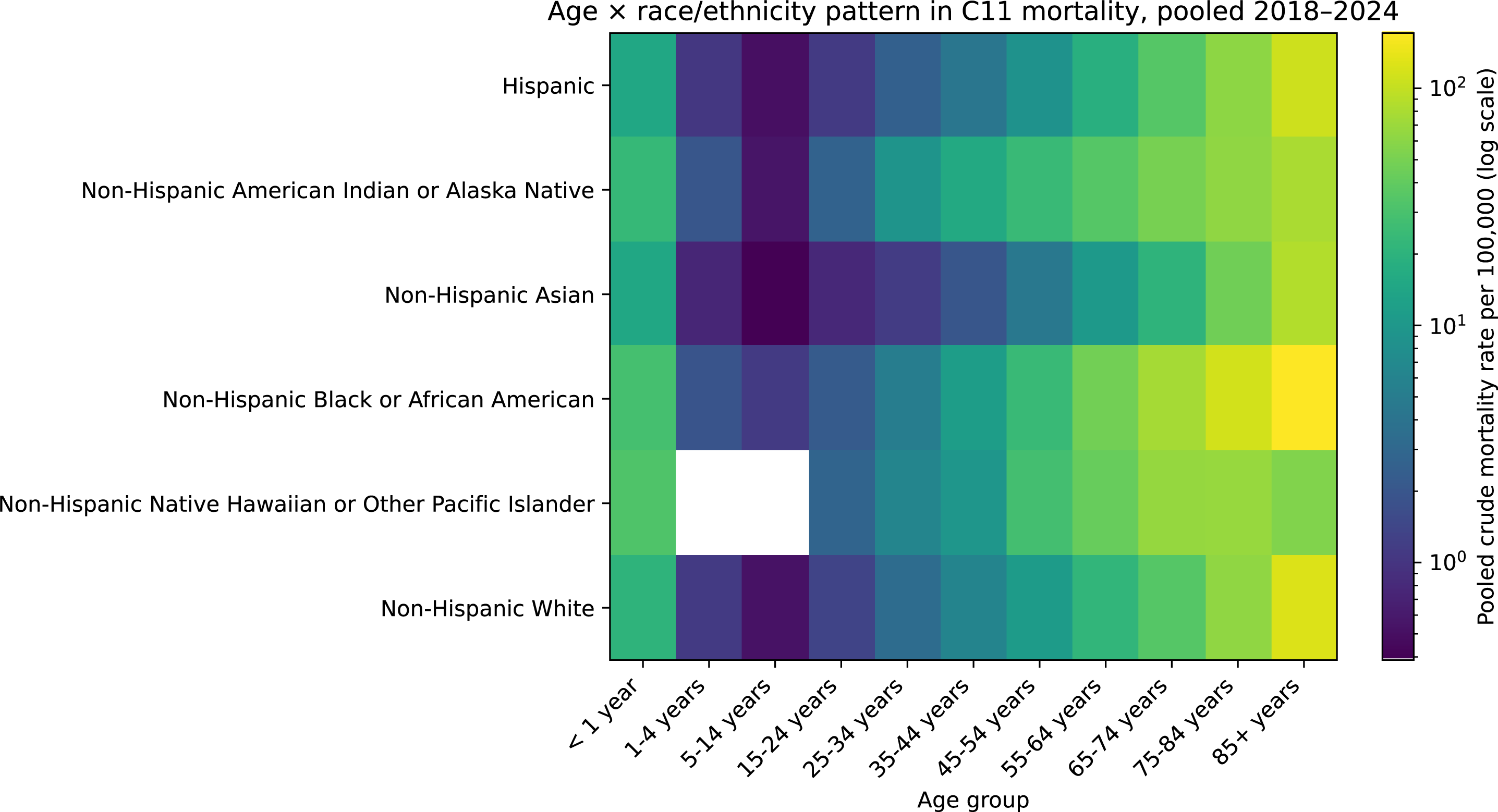

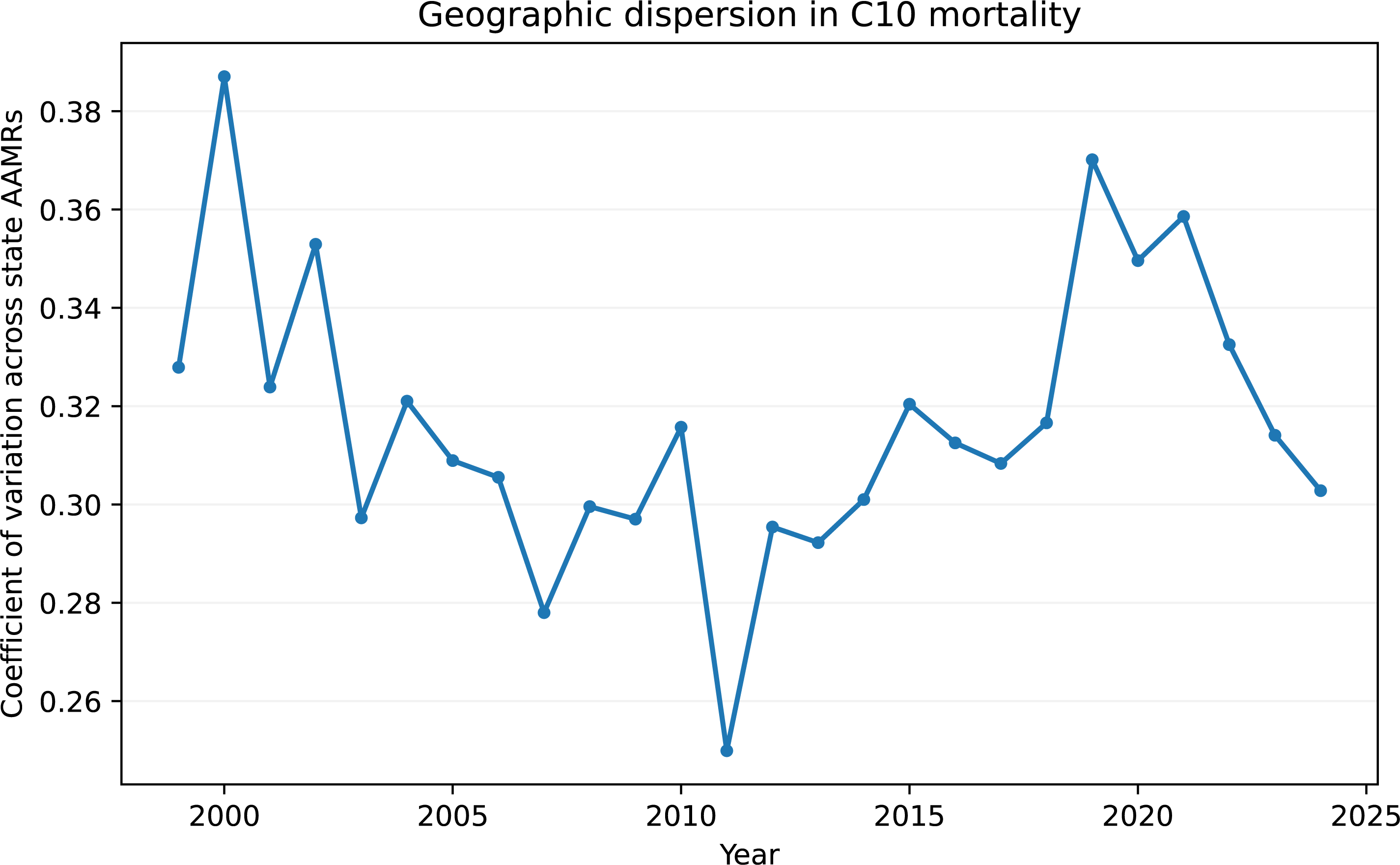

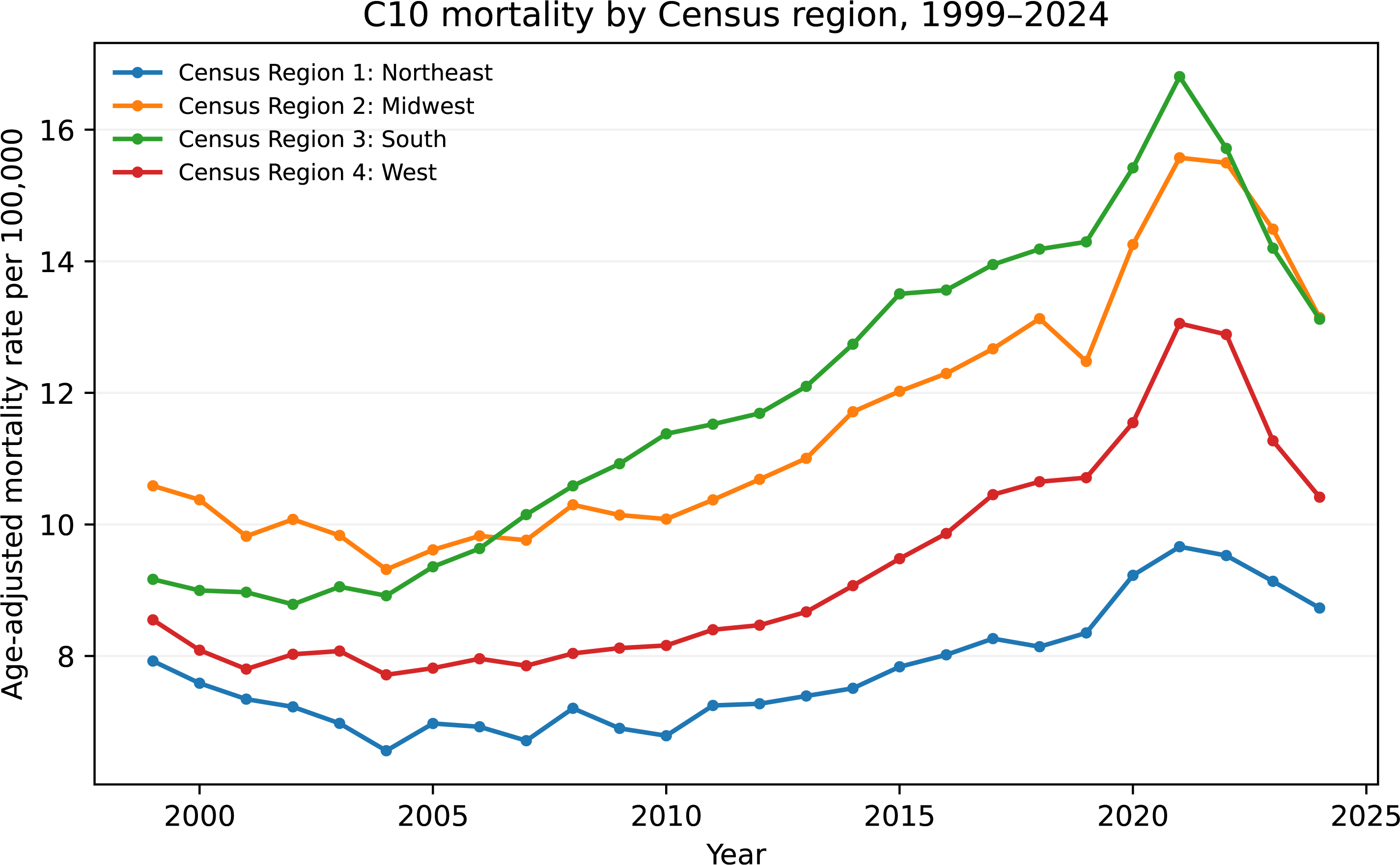

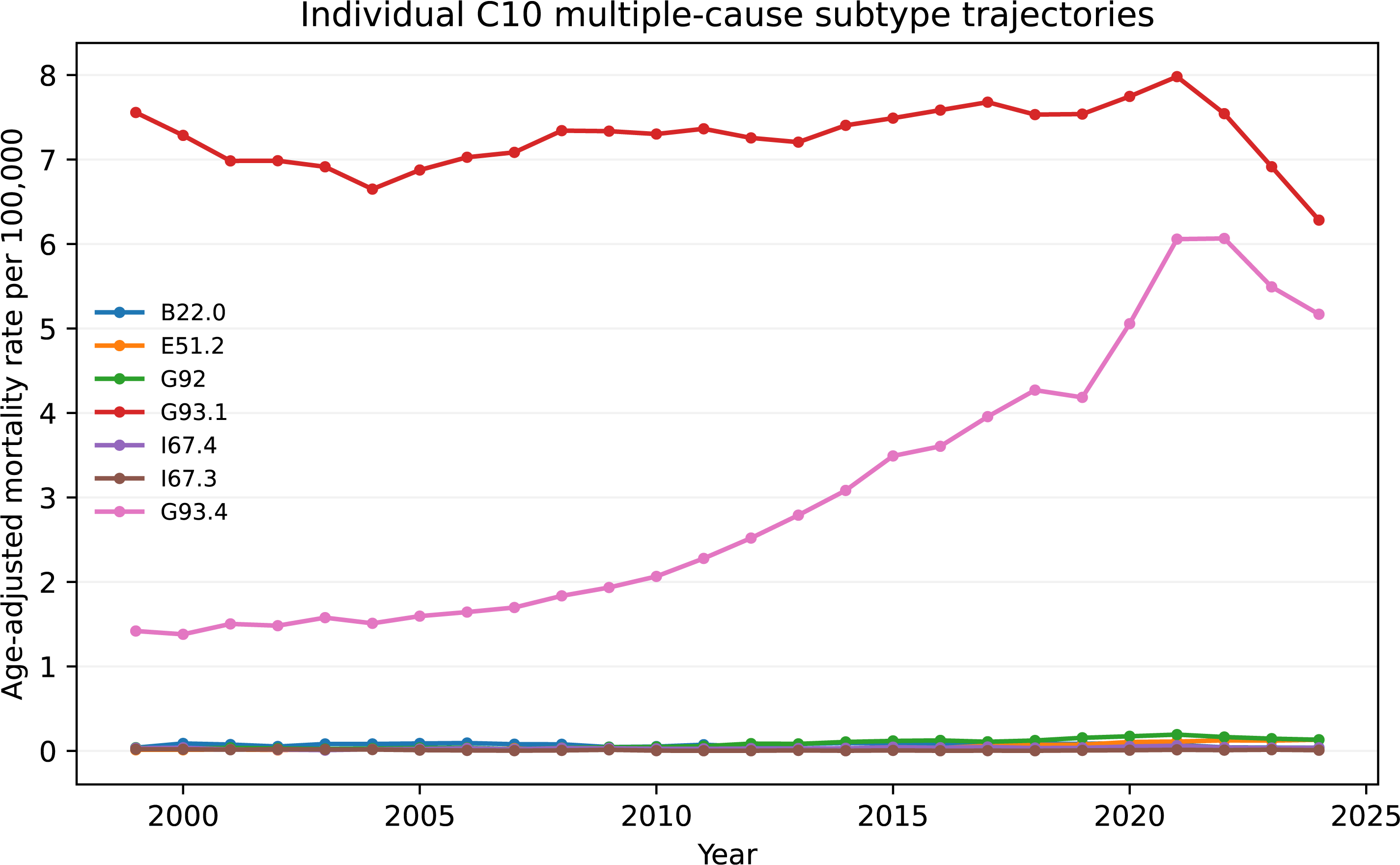

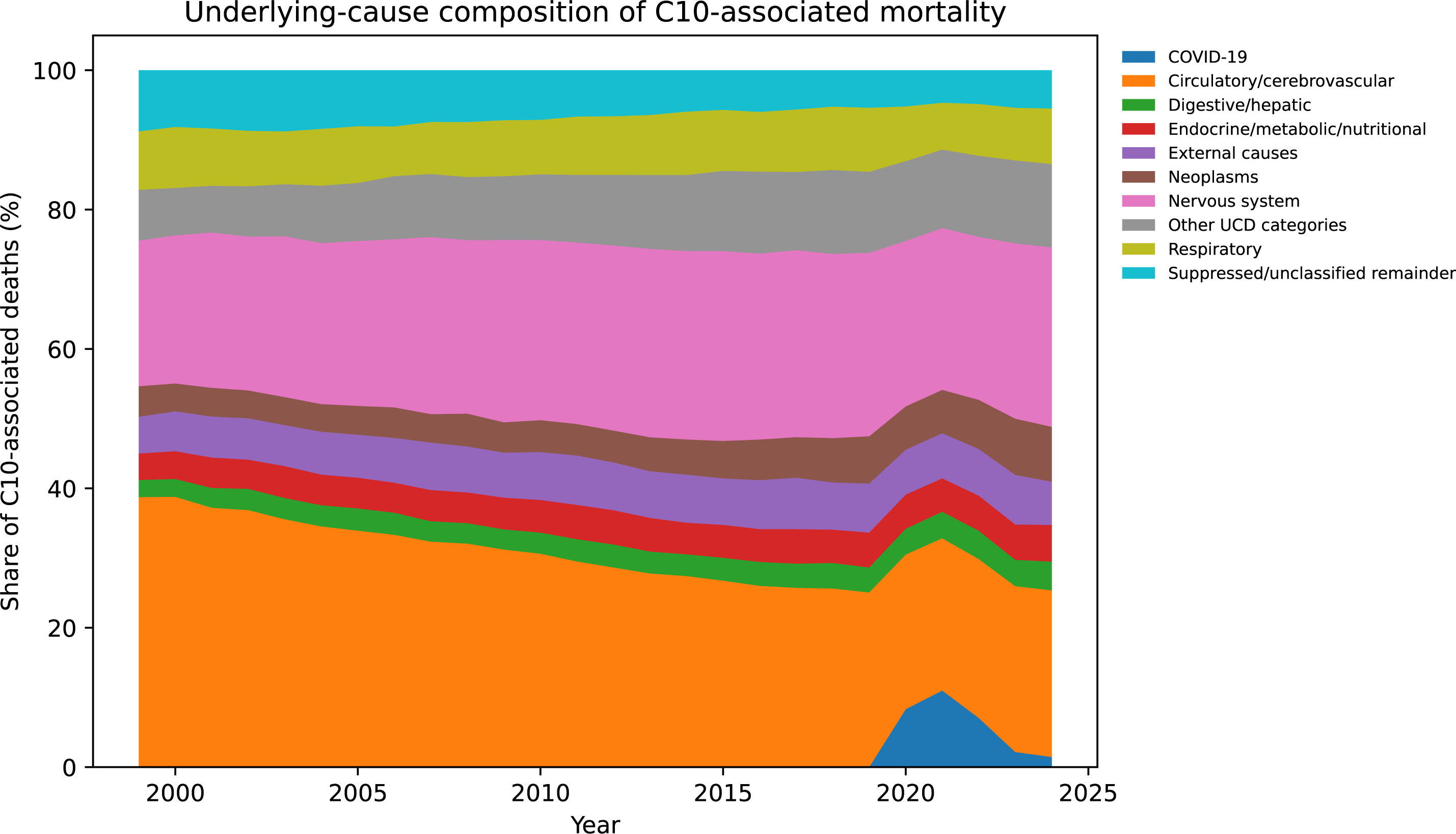

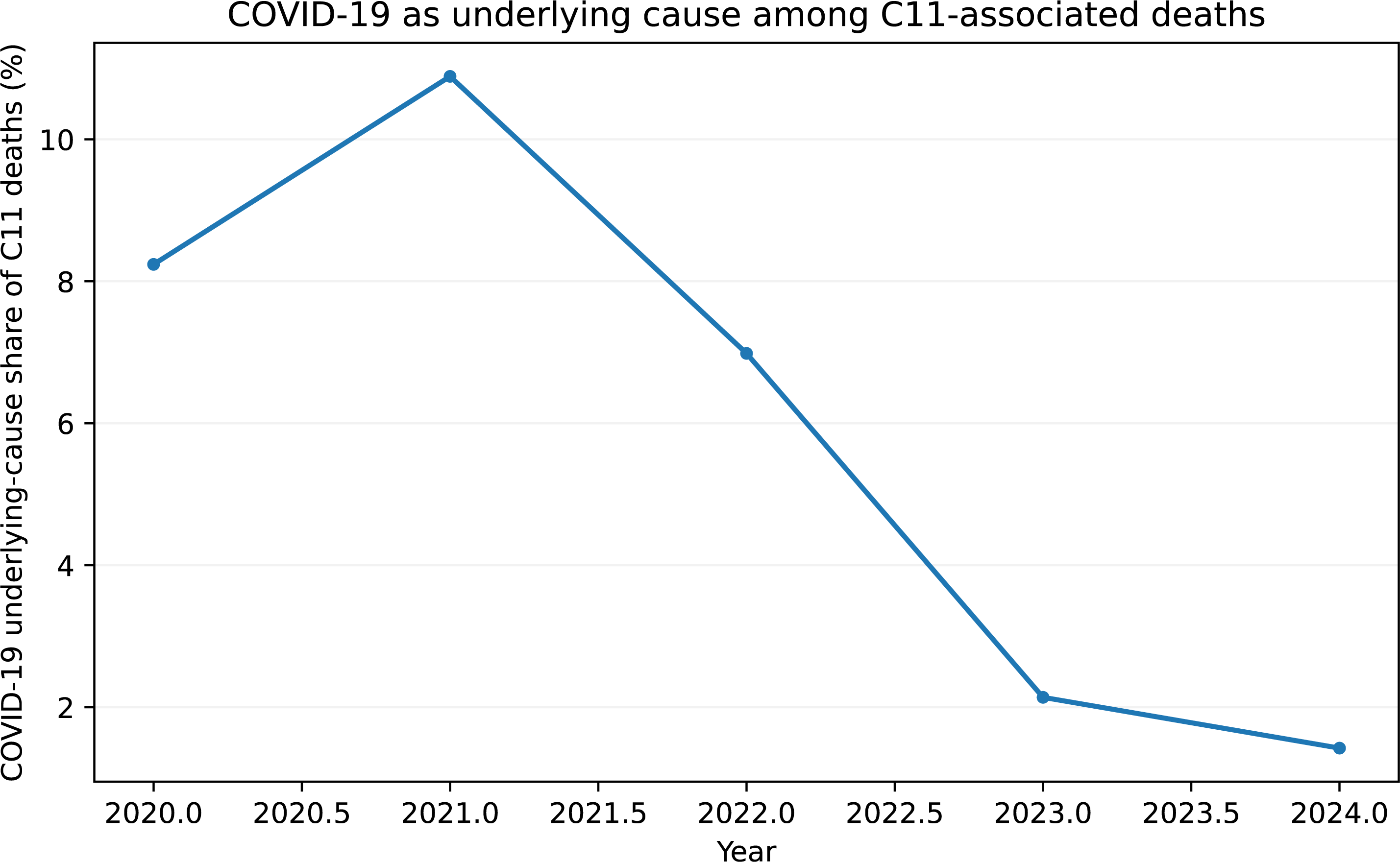

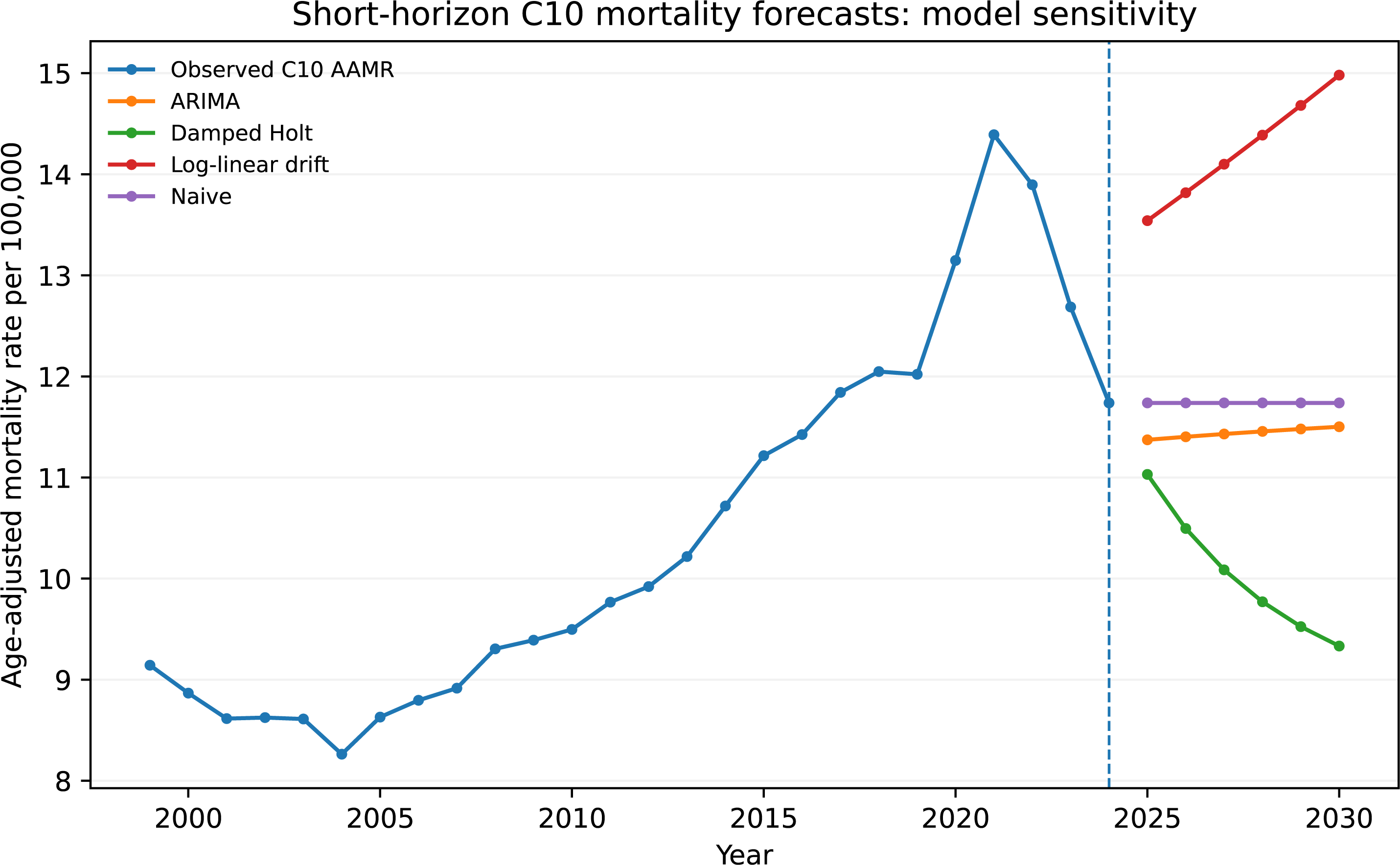

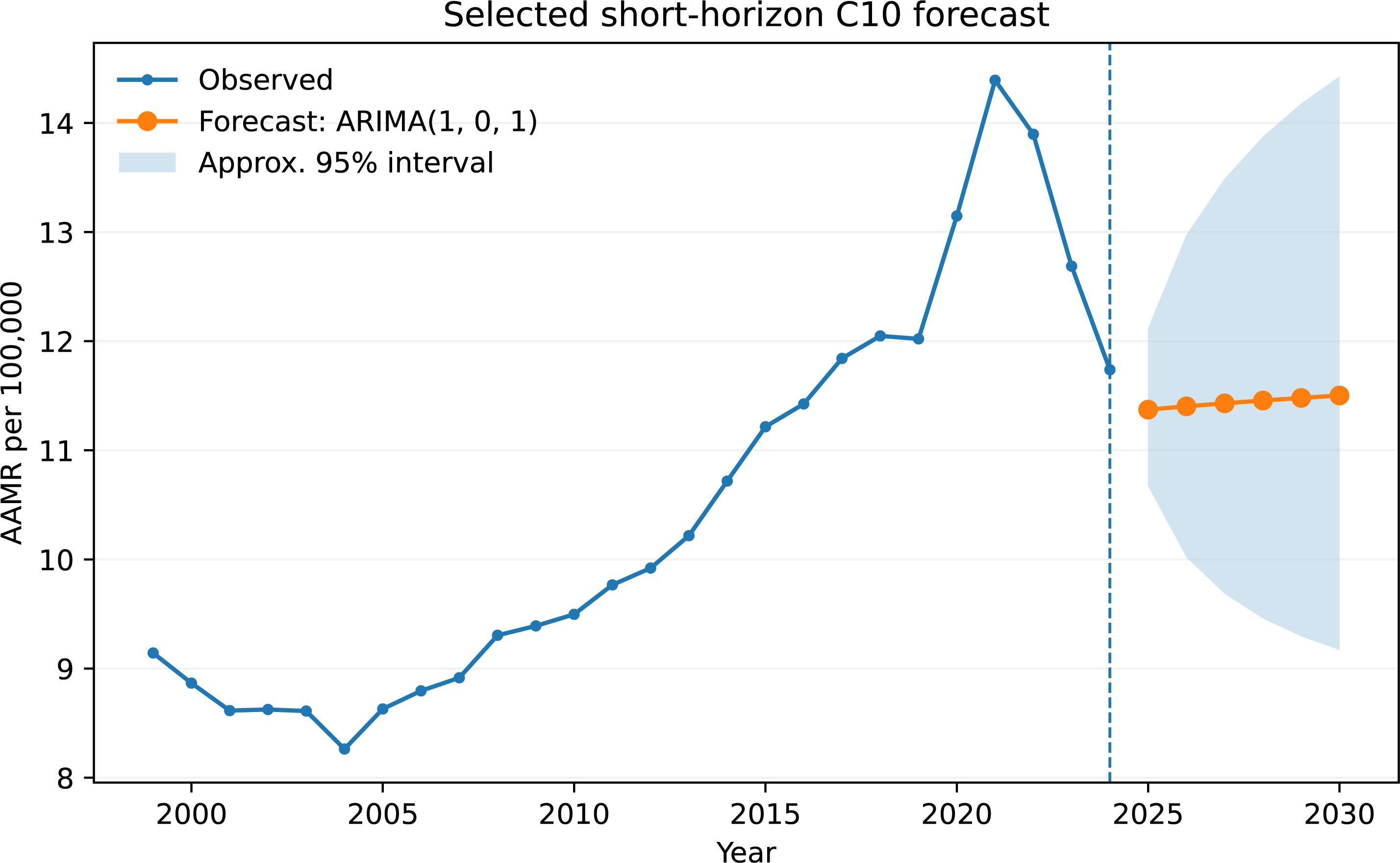

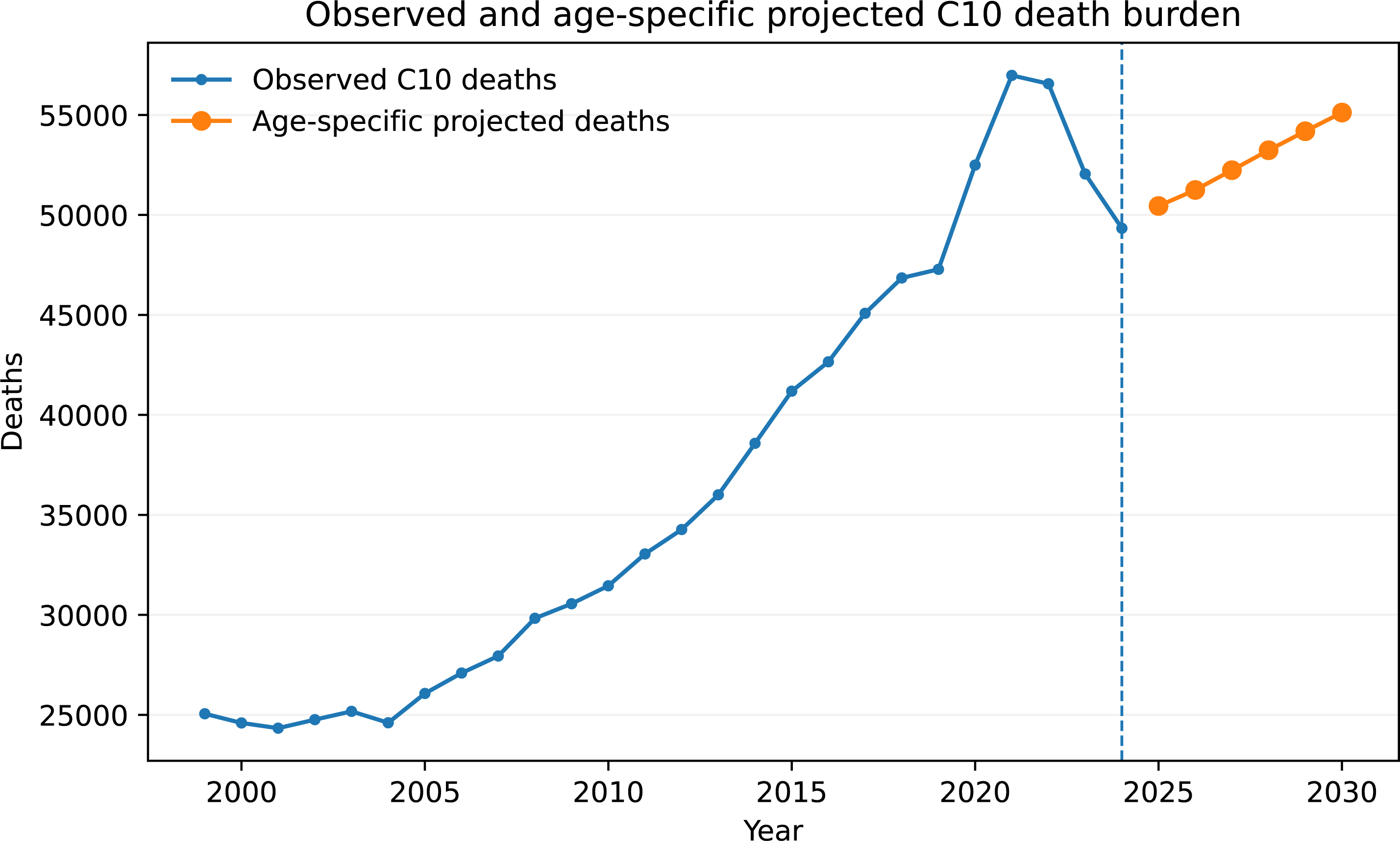

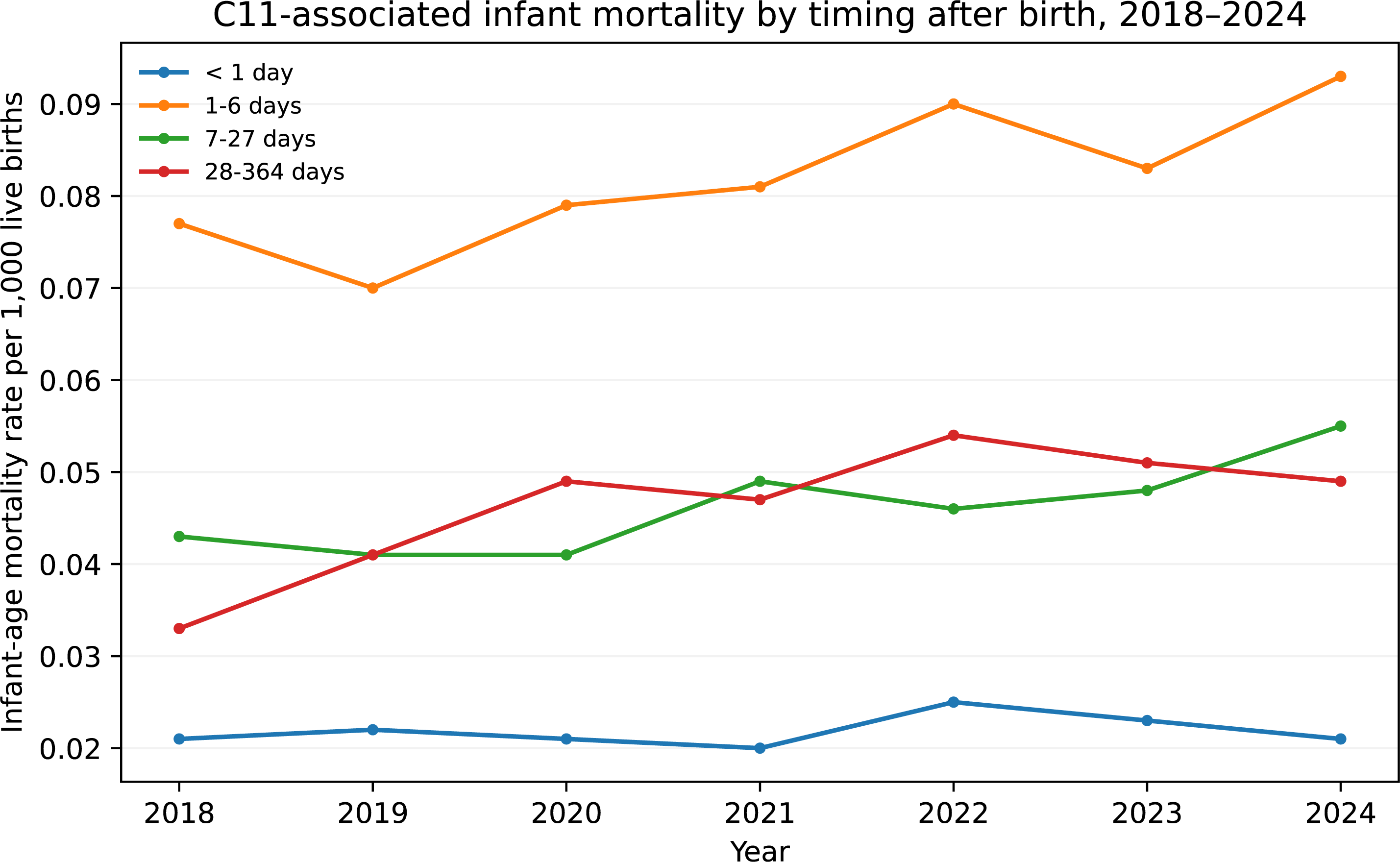

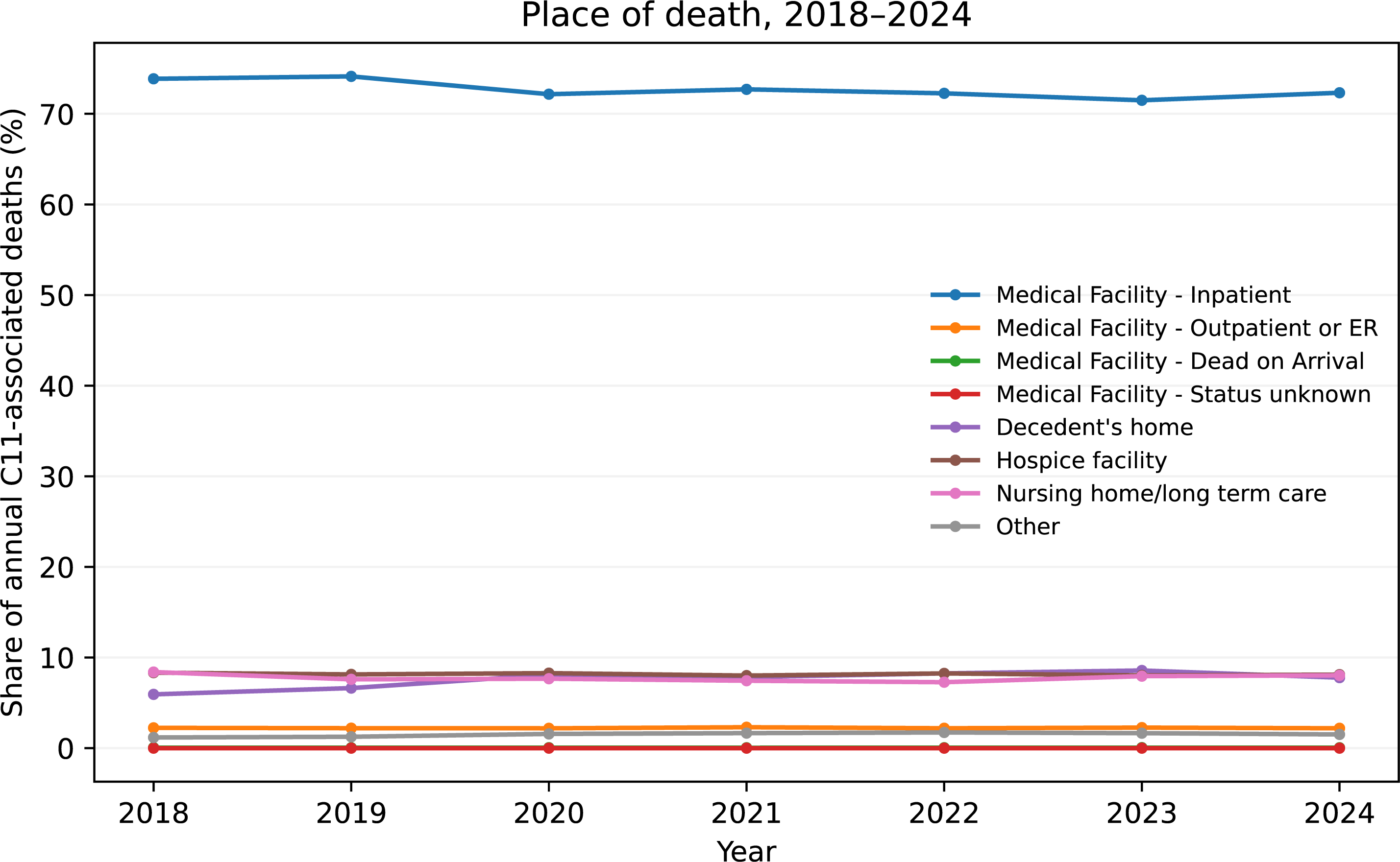

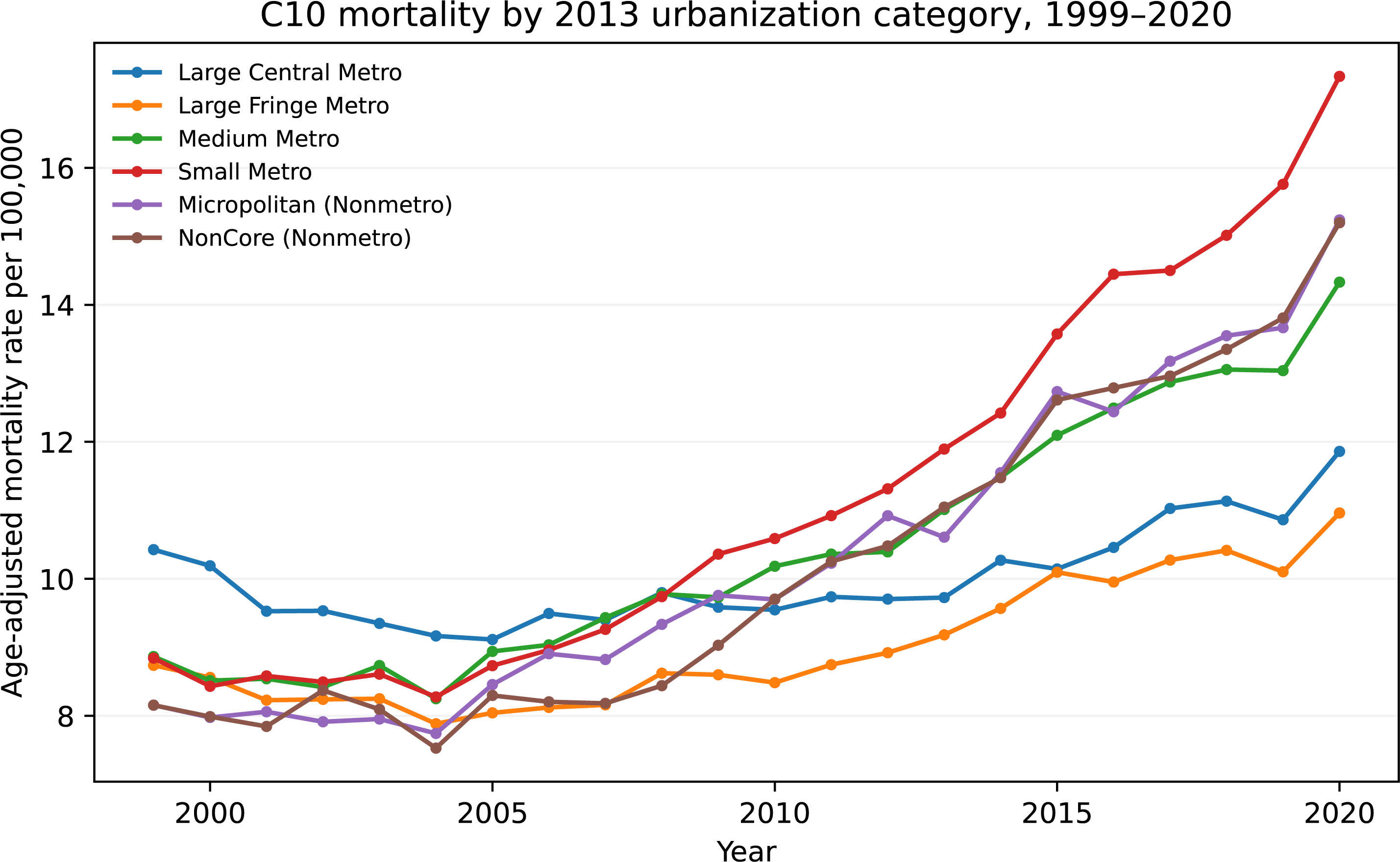

## References

1. Le Guennec L, Marois C, Demeret S, Wijdicks EFM, Weiss N. Toxic-metabolic encephalopathy in adults: critical discussion and pragmatical diagnostic approach. Rev Neurol (Paris). 2022;178:93–104. doi:10.1016/j.neurol.2021.11.007.

2. Steinberg A. Emergent management of hypoxic-ischemic brain injury. Continuum (Minneap Minn). 2024;30:588–610. doi:10.1212/CON.0000000000001426.

3. Sechi G, Serra A. Wernicke’s encephalopathy: new clinical settings and recent advances in diagnosis and management. Lancet Neurol. 2007;6:442–455. doi:10.1016/S1474-4422(07)70104-7.

4. Cantu-Weinstein A, Branning R, Alamir M, et al. Diagnosis and treatment of Wernicke’s encephalopathy: a systematic literature review. Gen Hosp Psychiatry. 2024;87:48–59. doi:10.1016/j.genhosppsych.2024.01.005.

5. Gewirtz AN, Gao V, Parauda SC, Robbins MS. Posterior reversible encephalopathy syndrome. Curr Pain Headache Rep. 2021;25:19. doi:10.1007/s11916-020-00932-1.

6. Acun C, Karnati S, Padiyar S, Puthuraya S, Aly H, Mohamed M. Trends of neonatal hypoxic-ischemic encephalopathy prevalence and associated risk factors in the United States, 2010 to 2018. Am J Obstet Gynecol. 2022;227:751.e1–751.e10. doi:10.1016/j.ajog.2022.06.002.

7. Centers for Disease Control and Prevention, National Center for Health Statistics. Multiple Cause of Death 1999-2020, CDC WONDER Online Database. https://wonder.cdc.gov/mcd-icd10.html. Accessed September 2026.

8. Centers for Disease Control and Prevention, National Center for Health Statistics. Multiple Cause of Death 2018-2024, Single Race, CDC WONDER Online Database. https://wonder.cdc.gov/mcd-icd10-expanded.html. Accessed September 2026.

9. Arias E, Heron M, Hakes J. The validity of race and Hispanic-origin reporting on death certificates in the United States: an update. Vital Health Stat 2. 2016;(172):1–21.

10. Alipour J, Payandeh A. Common errors in reporting cause-of-death statement on death certificates: a systematic review and meta-analysis. J Forensic Leg Med. 2021;82:102220. doi:10.1016/j.jflm.2021.102220.

11. Khan I, Ajaz IA, Chaudhary A, et al. Demographic and regional disparities in concurrent respiratory and renal failure mortality: a CDC WONDER analysis, 1999-2024. Sci Rep. 2026;16:24515. doi:10.1038/s41598-026-55835-9.

12. Javed H, Tariq S, Ahmer W, Afridi MK, Ahmed R. Trends and disparities in cerebral edema-related mortality in the United States: a nationwide analysis using CDC WONDER data, 1999-2023. J Clin Neurosci. 2026;146:111871. doi:10.1016/j.jocn.2026.111871.

13. von Elm E, Altman DG, Egger M, Pocock SJ, Gotzsche PC, Vandenbroucke JP; STROBE Initiative. The Strengthening the Reporting of Observational Studies in Epidemiology (STROBE) statement: guidelines for reporting observational studies. Lancet. 2007;370:1453–1457. doi:10.1016/S0140-6736(07)61602-X.

14. Anderson RN, Rosenberg HM. Age standardization of death rates: implementation of the year 2000 standard. Natl Vital Stat Rep. 1998;47(3):1–16,20.

15. Fay MP, Feuer EJ. Confidence intervals for directly standardized rates: a method based on the gamma distribution. Stat Med. 1997;16:791–801.

16. Kim HJ, Fay MP, Feuer EJ, Midthune DN. Permutation tests for joinpoint regression with applications to cancer rates. Stat Med. 2000;19:335–351.

17. Mao L, Jin H, Wang M, et al. Neurologic manifestations of hospitalized patients with coronavirus disease 2019 in Wuhan, China. JAMA Neurol. 2020;77:683–690. doi:10.1001/jamaneurol.2020.1127.

18. Misra S, Kolappa K, Prasad M, et al. Frequency of neurologic manifestations in COVID-19: a systematic review and meta-analysis. Neurology. 2021;97:e2269–e2281. doi:10.1212/WNL.0000000000012930.

