## Supplementary Information for "National trends, demographic disparities, and phenotypic shifts in encephalopathy-associated mortality in the United States: A CDC WONDER analysis, 1999-2024"

### Supplementary Methods

#### Phenotype definitions

The primary C10 phenotype was designed as a time-stable syndrome definition for 1999-2024. C11 adds neonatal hypoxic-ischemic encephalopathy (P91.6) and is used from 2006 onward. G93.4 provides a narrow unspecified-encephalopathy sensitivity definition. B14 and B15 broaden the syndrome definition to test robustness to inclusion of additional encephalopathic disorders.

#### Supplementary Table 1 | Encephalopathy phenotype definitions.

| **Phenotype** | **Years** | **ICD-10 definition** | **Analytic role** |
| --- | --- | --- | --- |
| C10 | 1999-2024 | B22.0, E51.2, G92, G93.1, G93.4, I67.3, I67.4, P57.0, P57.8, P57.9 | Primary time-stable multiple-cause phenotype. |
| C11 | 2006-2024 | C10 + P91.6 | Comprehensive phenotype including neonatal hypoxic-ischemic encephalopathy. |
| G93.4 | 1999-2024 | G93.4 | Narrow sensitivity definition: encephalopathy, unspecified. |
| B14 | 1999-2024 | C10 + A81.0, A81.1, A81.2, G93.7 | Broader sensitivity definition. |
| B15 | 2006-2024 | B14 + P91.6 | Broadest sensitivity definition used from 2006 onward. |

**Deaths were identified when at least one listed phenotype code appeared among the multiple causes unless an underlying-cause restriction was explicitly applied.**

#### Database overlap validation

To validate the transition between CDC WONDER data products, national C10 and C11 estimates were compared in the 2018-2020 overlap. Counts, populations, crude rates, and AAMRs were identical in every overlap year. For C10, the only audited discrepancy was an AAMR standard error of 0.057 versus 0.056 in 2019.

#### Supplementary Table 2 | National overlap validation across CDC WONDER databases, 2018-2020.

| **Phenotype** | **Year** | **Deaths** | **AAMR, D77** | **AAMR, D157** | **SE, D77** | **SE, D157** |
| --- | --- | --- | --- | --- | --- | --- |
| C10 | 2018 | 46,850 | 12.048 | 12.048 | 0.057 | 0.057 |
| C10 | 2019 | 47,280 | 12.021 | 12.021 | 0.057 | 0.056 |
| C10 | 2020 | 52,494 | 13.147 | 13.147 | 0.059 | 0.059 |
| C11 | 2018 | 47,344 | 12.223 | 12.223 | 0.057 | 0.057 |
| C11 | 2019 | 47,740 | 12.190 | 12.190 | 0.057 | 0.057 |
| C11 | 2020 | 52,975 | 13.340 | 13.340 | 0.059 | 0.059 |

**AAMR, age-adjusted mortality rate per 100,000; SE, standard error. Death counts and population denominators were identical across databases for all rows.**

### Supplementary Results

#### Infant-age strata

#### Supplementary Table 3 | C11-associated infant mortality rate by timing after birth, 2018-2024.

| **Infant age** | **2018** | **2019** | **2020** | **2021** | **2022** | **2023** | **2024** |
| --- | --- | --- | --- | --- | --- | --- | --- |
| < 1 day | 0.021 | 0.022 | 0.021 | 0.020 | 0.025 | 0.023 | 0.021 |
| 1-6 days | 0.077 | 0.070 | 0.079 | 0.081 | 0.090 | 0.083 | 0.093 |
| 7-27 days | 0.043 | 0.041 | 0.041 | 0.049 | 0.046 | 0.048 | 0.055 |
| 28-364 days | 0.033 | 0.041 | 0.049 | 0.047 | 0.054 | 0.051 | 0.049 |

**Rates are deaths per 1,000 live births, matching the supplied infant-age analysis output.**

#### Supplementary Table 4 | Under-1-year perinatal encephalopathy component deaths, 2018-2024.

| **Code** | **Diagnosis** | **2018** | **2019** | **2020** | **2021** | **2022** | **2023** | **2024** |
| --- | --- | --- | --- | --- | --- | --- | --- | --- |
| P57.0 | Kernicterus due to isoimmunization | 0 | Suppressed | 0 | 0 | 0 | 0 | 0 |
| P57.8 | Other specified kernicterus | 0 | 0 | 0 | 0 | 0 | 0 | 0 |
| P57.9 | Kernicterus, unspecified | 0 | 0 | 0 | Suppressed | Suppressed | 0 | 0 |
| P91.6 | Hypoxic ischemic encephalopathy of newborn | 473 | 452 | 470 | 497 | 544 | 536 | 547 |

**Counts are shown as extracted. Suppressed cells remain suppressed and were not imputed as zero.**

#### Place of death, autopsy, weekday, month, and education

#### Supplementary Table 5 | Place-of-death distribution among C11-associated deaths, 2018-2024 (%).

| **Place of death** | **2018** | **2019** | **2020** | **2021** | **2022** | **2023** | **2024** |
| --- | --- | --- | --- | --- | --- | --- | --- |
| Medical Facility - Inpatient | 73.86 | 74.13 | 72.16 | 72.70 | 72.26 | 71.49 | 72.32 |
| Medical Facility - Outpatient or ER | 2.24 | 2.20 | 2.19 | 2.31 | 2.20 | 2.26 | 2.19 |
| Medical Facility - Dead on Arrival | 0.05 | 0.04 | 0.05 | 0.04 | 0.04 | 0.05 | 0.04 |
| Medical Facility - Status unknown | 0.00 | 0.00 | 0.00 | 0.00 | 0.00 | 0.00 | 0.00 |
| Decedent's home | 5.93 | 6.63 | 8.09 | 7.85 | 8.25 | 8.57 | 7.78 |
| Hospice facility | 8.33 | 8.14 | 8.28 | 8.01 | 8.25 | 8.03 | 8.11 |
| Nursing home/long term care | 8.40 | 7.59 | 7.66 | 7.44 | 7.28 | 7.94 | 8.03 |
| Other | 1.17 | 1.25 | 1.57 | 1.66 | 1.73 | 1.65 | 1.51 |

**Values are percentages of annual C11-associated deaths.**

#### Supplementary Table 6 | Autopsy-status distribution among C11-associated deaths, 2018-2024 (%).

| **Autopsy status** | **2018** | **2019** | **2020** | **2021** | **2022** | **2023** | **2024** |
| --- | --- | --- | --- | --- | --- | --- | --- |
| No | 92.81 | 92.72 | 93.83 | 93.65 | 94.02 | 93.64 | 93.32 |
| Yes | 3.66 | 3.78 | 3.00 | 2.85 | 2.75 | 2.68 | 2.52 |
| Unknown | 3.52 | 3.50 | 3.17 | 3.49 | 3.23 | 3.67 | 4.16 |

**Values are percentages of annual C11-associated deaths.**

#### Supplementary Table 7 | Weekday distribution of C10-associated deaths, 2018-2024 (%).

| **Weekday** | **2018** | **2019** | **2020** | **2021** | **2022** | **2023** | **2024** |
| --- | --- | --- | --- | --- | --- | --- | --- |
| Sunday | 13.25 | 13.55 | 13.42 | 13.37 | 13.43 | 13.91 | 13.70 |
| Monday | 14.19 | 14.04 | 13.83 | 13.63 | 13.56 | 14.14 | 14.07 |
| Tuesday | 14.70 | 14.57 | 14.34 | 14.48 | 14.33 | 14.13 | 14.34 |
| Wednesday | 14.59 | 14.53 | 14.66 | 14.56 | 14.81 | 14.52 | 14.26 |
| Thursday | 14.48 | 14.49 | 14.85 | 14.48 | 14.38 | 14.45 | 14.41 |
| Friday | 14.85 | 14.69 | 14.71 | 15.19 | 14.98 | 14.66 | 14.98 |
| Saturday | 13.94 | 14.12 | 14.20 | 14.28 | 14.52 | 14.20 | 14.23 |
| Unknown | 0.00 | 0.00 | 0.00 | 0.00 | 0.00 | 0.00 | 0.00 |

**Values are percentages of annual C10-associated deaths.**

#### Supplementary Table 8 | Monthly distribution of C10-associated deaths, 2018-2024 (%).

| **Month** | **2018** | **2019** | **2020** | **2021** | **2022** | **2023** | **2024** |
| --- | --- | --- | --- | --- | --- | --- | --- |
| Jan | 10.36 | 9.29 | 8.27 | 10.41 | 10.98 | 9.84 | 10.07 |
| Feb | 8.42 | 8.23 | 7.53 | 8.29 | 9.18 | 8.07 | 8.64 |
| Mar | 8.79 | 9.12 | 7.74 | 8.39 | 8.59 | 9.04 | 8.87 |
| Apr | 8.14 | 8.58 | 7.53 | 7.64 | 7.82 | 8.45 | 8.28 |
| May | 8.07 | 8.44 | 8.05 | 7.70 | 7.69 | 8.35 | 8.26 |
| Jun | 7.61 | 7.92 | 7.73 | 7.18 | 7.55 | 7.60 | 8.00 |
| Jul | 7.69 | 7.84 | 8.44 | 7.47 | 7.98 | 7.54 | 7.58 |
| Aug | 7.87 | 7.83 | 8.21 | 8.23 | 7.85 | 7.98 | 7.74 |
| Sep | 7.64 | 7.39 | 7.94 | 8.46 | 7.49 | 7.71 | 7.74 |
| Oct | 8.19 | 8.05 | 8.58 | 8.44 | 7.92 | 8.16 | 8.21 |
| Nov | 8.44 | 8.27 | 9.19 | 8.61 | 7.79 | 8.18 | 7.89 |
| Dec | 8.77 | 9.04 | 10.80 | 9.20 | 9.18 | 9.06 | 8.72 |

**Values are percentages of annual C10-associated deaths.**

#### Supplementary Table 9 | Education distribution among C11-associated deaths, 2021-2024 (%).

| **Education** | **2021** | **2022** | **2023** | **2024** |
| --- | --- | --- | --- | --- |
| 8th grade or less | 9.36 | 8.99 | 8.82 | 8.85 |
| 9th through 12th grade with no diploma | 10.36 | 10.41 | 9.76 | 9.58 |
| High school graduate or GED completed | 42.32 | 42.16 | 41.85 | 41.47 |
| Some college credit, but not a degree | 13.31 | 13.05 | 13.21 | 13.16 |
| Associate degree (AA,AS) | 6.99 | 6.88 | 7.07 | 7.24 |
| Bachelor's degree (BA, AB, BS) | 10.06 | 10.38 | 10.71 | 10.89 |
| Master's degree (MA, MS, MEng, MEd, MSW, MBA) | 3.86 | 4.14 | 4.44 | 4.55 |
| Doctorate (PhD, EdD) or Professional Degree (MD, DDS, DVM, LLB, JD) | 1.52 | 1.65 | 1.72 | 1.81 |
| Unknown or Not Stated | 2.22 | 2.33 | 2.41 | 2.45 |
| Not Available | 0.00 | 0.00 | 0.00 | 0.00 |

**Values are percentages of annual C11-associated deaths. Education is descriptive because population denominators were not applicable in the supplied extract.**

#### Subtype and COVID-19 sensitivity analyses

#### Supplementary Table 10 | Exploratory long-term trends for selected individual C10 component codes.

| **Code** | **Label** | **Years, n** | **APC, %** | **95% CI** | **P value** | **BH q value** |
| --- | --- | --- | --- | --- | --- | --- |
| B22.0 | HIV disease resulting in encephalopathy | 26 | -0.57 | -2.08 to 0.96 | 0.472 | 0.472 |
| E51.2 | Wernicke encephalopathy | 26 | 12.34 | 10.64 to 14.06 | 1.14e-13 | 3.98e-13 |
| G92 | Toxic encephalopathy | 26 | 8.85 | 7.21 to 10.52 | 8.64e-11 | 2.02e-10 |
| G93.1 | Anoxic brain damage, not elsewhere classified | 26 | 0.18 | -0.09 to 0.45 | 0.202 | 0.236 |
| G93.4 | Encephalopathy, unspecified | 26 | 7.11 | 6.43 to 7.79 | 5e-17 | 3.5e-16 |
| I67.3 | Progressive vascular leukoencephalopathy | 26 | -2.48 | -4.97 to 0.07 | 0.0686 | 0.096 |
| I67.4 | Hypertensive encephalopathy | 26 | 2.83 | 1.40 to 4.28 | 0.000653 | 0.00114 |

**APC, annual percent change; BH, Benjamini-Hochberg false-discovery-rate adjustment. These code-specific trends are exploratory.**

#### Supplementary Table 11 | COVID-19 as the underlying cause among C11-associated deaths, 2020-2024.

| **Year** | **COVID-19 underlying-cause deaths** | **All C11-associated deaths** | **COVID-19 share, %** |
| --- | --- | --- | --- |
| 2020 | 4,364 | 52,975 | 8.24 |
| 2021 | 6,259 | 57,494 | 10.89 |
| 2022 | 3,989 | 57,116 | 6.98 |
| 2023 | 1,125 | 52,590 | 2.14 |
| 2024 | 710 | 49,882 | 1.42 |

**U07.1 was used to identify COVID-19 as the underlying cause.**

#### Exploratory forecasting

#### Supplementary Table 12 | Rolling-origin forecast model comparison.

| **Model** | **Forecasts, n** | **MAE** | **RMSE** | **MASE** |
| --- | --- | --- | --- | --- |
| ARIMA | 14 | 0.529 | 0.791 | 1.357 |
| Naive | 14 | 0.543 | 0.674 | 1.394 |
| Damped Holt | 14 | 0.613 | 0.854 | 1.574 |
| Log-linear drift | 14 | 0.935 | 1.044 | 2.400 |

**All models had MASE >1 in the supplied validation output. Forecasting is exploratory and not part of the primary inference.**

#### Supplementary Table 13 | Selected national C10 AAMR forecast scenario, 2025-2030.

| **Year** | **Model** | **Forecast AAMR** | **Lower 95%** | **Upper 95%** |
| --- | --- | --- | --- | --- |
| 2025 | ARIMA(1, 0, 1) | 11.373 | 10.671 | 12.121 |
| 2026 | ARIMA(1, 0, 1) | 11.403 | 10.022 | 12.974 |
| 2027 | ARIMA(1, 0, 1) | 11.430 | 9.685 | 13.490 |
| 2028 | ARIMA(1, 0, 1) | 11.456 | 9.460 | 13.874 |
| 2029 | ARIMA(1, 0, 1) | 11.480 | 9.296 | 14.178 |
| 2030 | ARIMA(1, 0, 1) | 11.502 | 9.171 | 14.425 |

**Rates are per 100,000. Intervals are model-based approximate 95% intervals from the supplied exploratory forecast output.**

#### Supplementary Table 14 | Age-specific projected C10 death burden, 2025-2030.

| **Year** | **Projected deaths** |
| --- | --- |
| 2025 | 50,448 |
| 2026 | 51,248 |
| 2027 | 52,238 |
| 2028 | 53,236 |
| 2029 | 54,186 |
| 2030 | 55,120 |

**Projected totals are generated from the supplied age-specific rate and population scenario and should be interpreted as conditional scenarios.**

#### Supplementary Table 15 | Decomposition of projected C10 death burden into population-composition and rate effects.

| **Year** | **Baseline deaths** | **Projected deaths** | **Population effect** | **Rate effect** |
| --- | --- | --- | --- | --- |
| 2025 | 49,333 | 50,448 | 1,316 | -200 |
| 2026 | 49,333 | 51,248 | 2,274 | -359 |
| 2027 | 49,333 | 52,238 | 3,383 | -477 |
| 2028 | 49,333 | 53,236 | 4,469 | -565 |
| 2029 | 49,333 | 54,186 | 5,486 | -633 |
| 2030 | 49,333 | 55,120 | 6,474 | -687 |

**The decomposition follows the supplied age-specific forecasting output.**

#### Supplementary Table 16 | Final analytic quality-control checks.

| **Check** | **Status** | **Detail** |
| --- | --- | --- |
| C10 spans 1999–2024 | Pass | (1999, 2024) |
| C10 has one national row per year | Pass | n=26 |
| C11 begins 2006 | Pass | (2006, 2024) |
| No imputed suppressed C10 national deaths | Pass | — |
| C10 overlap death differences small | Pass | 0 0.0 1 0.0 2 0.0 Name: deaths_diff, dtype: float64 |
| C11 overlap death differences small | Pass | 0 0.0 1 0.0 2 0.0 Name: deaths_diff, dtype: float64 |
| State map includes 2024 | Pass | (1999, 2024) |
| Forecast horizon ends at configured year | Pass | 2030 |

**Quality-control checks are reproduced from the supplied analysis output.**

### Supplementary Figures

#### Supplementary Figure 1 | C11-associated infant mortality by timing after birth, 2018-2024.

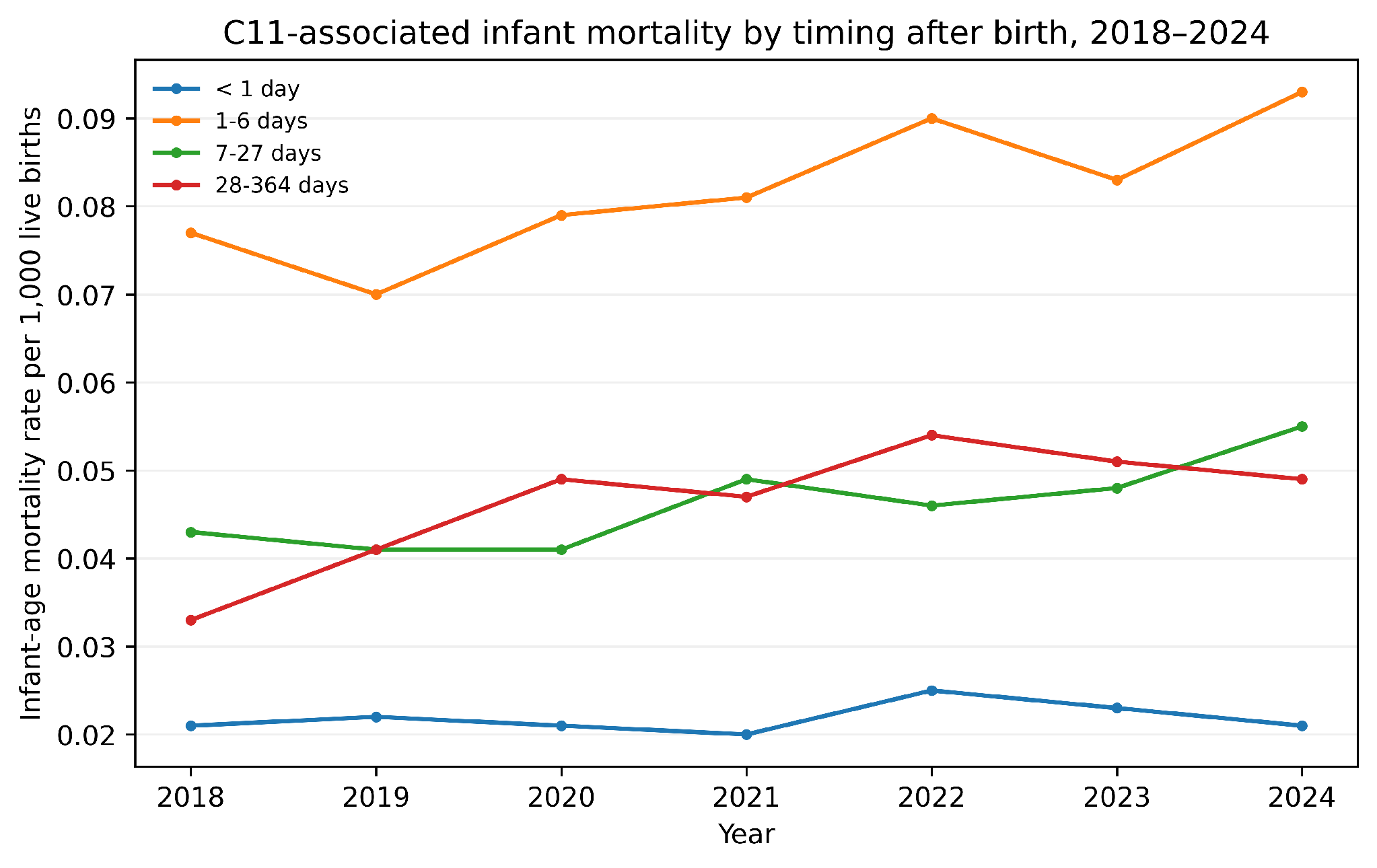

Rates are deaths per 1,000 live births across four infant-age strata.

#### Supplementary Figure 2 | C10 mortality by 2013 urbanization category, 1999-2020.

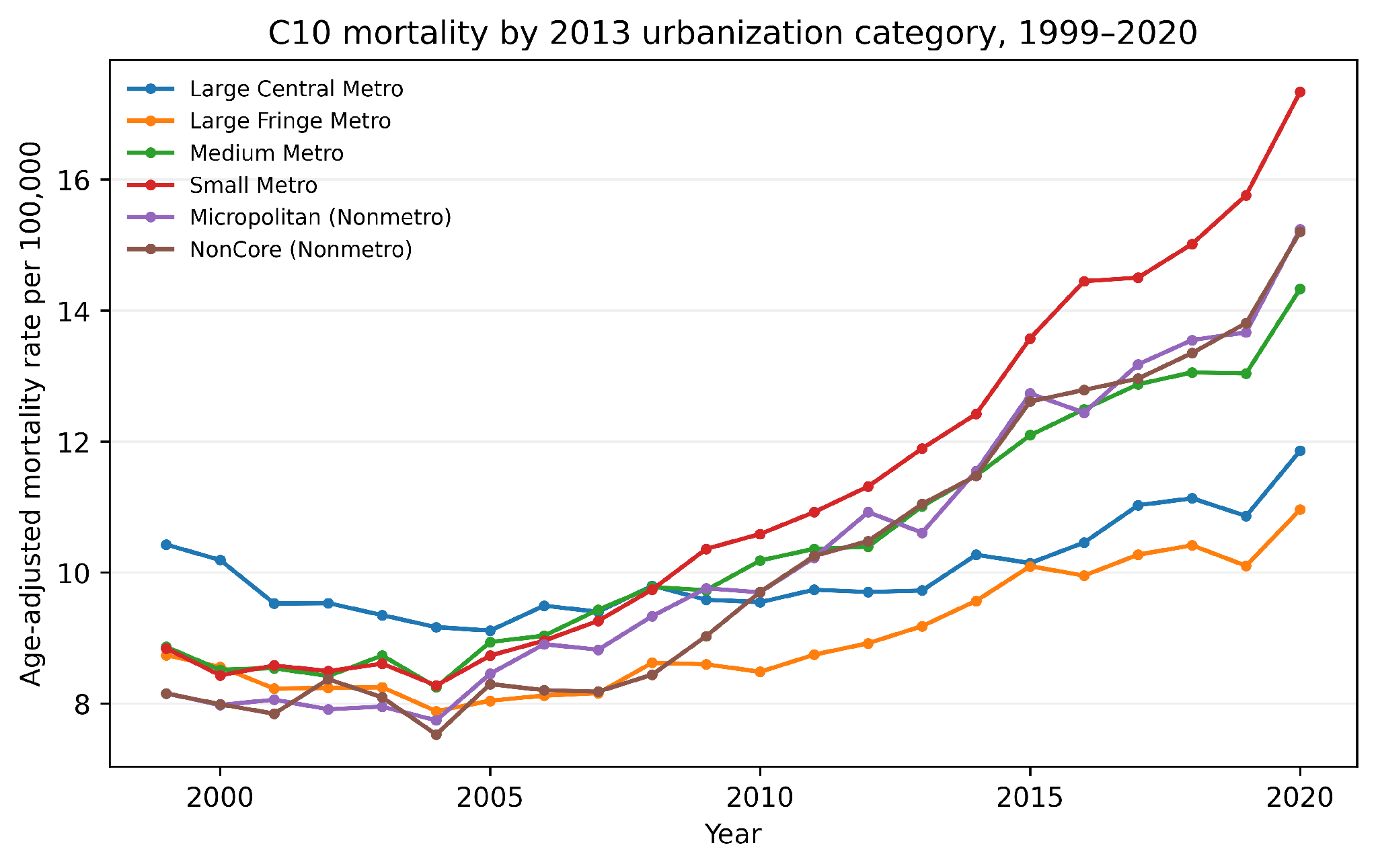

Age-adjusted mortality rates are shown per 100,000 for six urban-rural categories.

#### Supplementary Figure 3 | Place-of-death distribution among C11-associated deaths, 2018-2024.

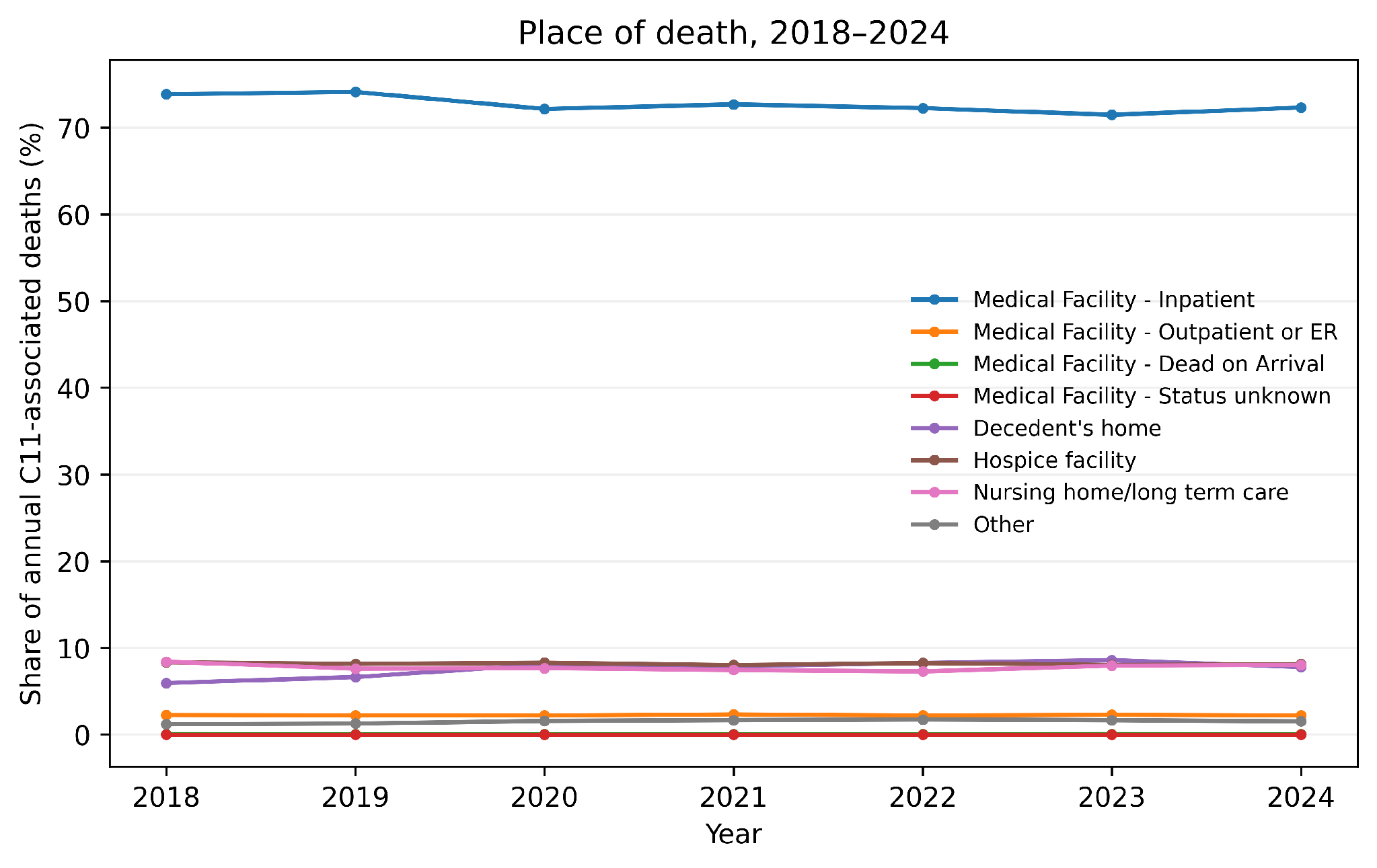

Lines show the annual share of C11-associated deaths across place-of-death categories.

#### Supplementary Figure 4 | Short-horizon C10 mortality forecast model sensitivity.

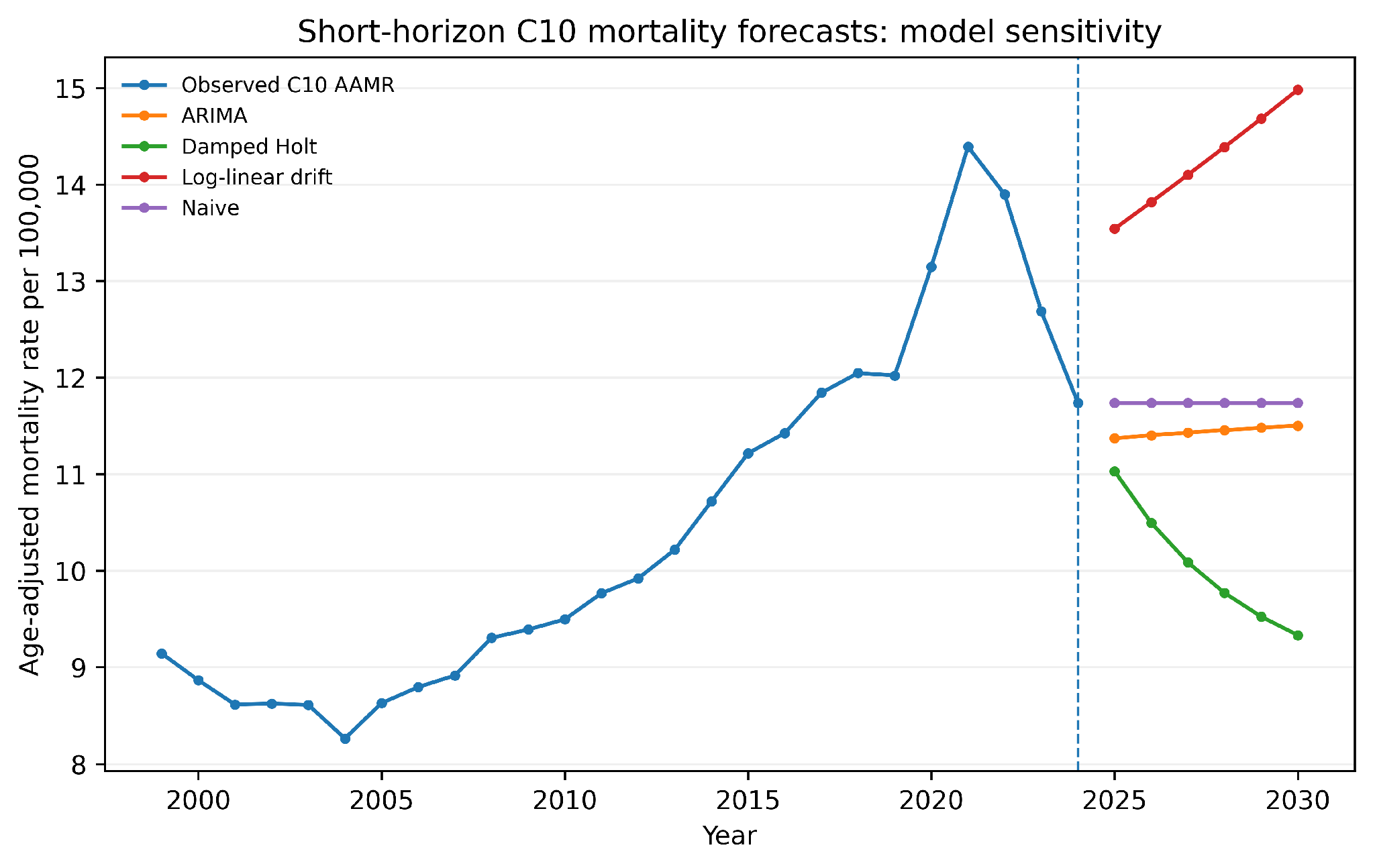

Observed national C10 AAMR is shown with exploratory forecasts from ARIMA, damped Holt, log-linear drift, and naive models for 2025-2030.

#### Supplementary Figure 5 | Selected short-horizon C10 forecast.

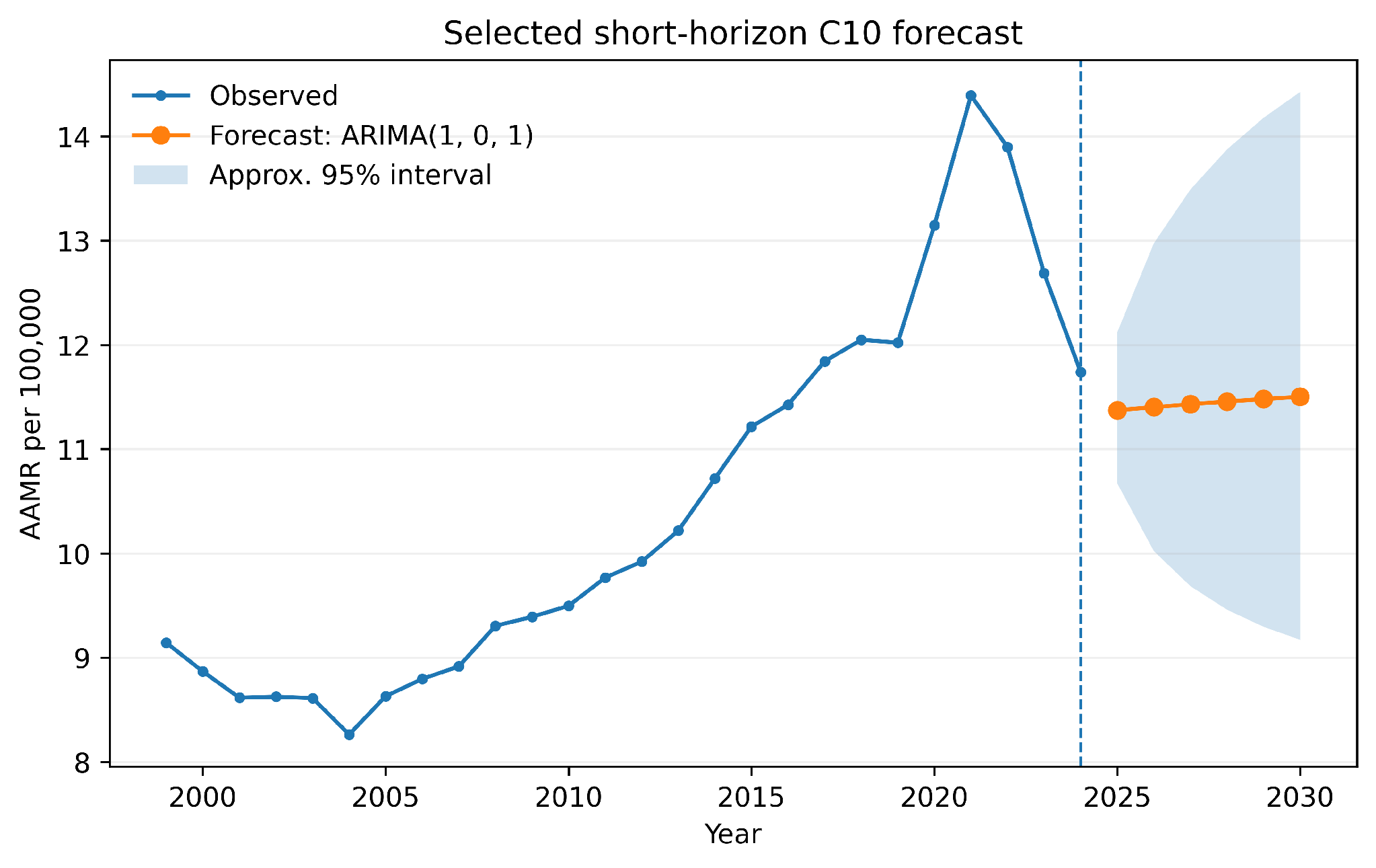

Observed C10 AAMR is shown with the selected ARIMA(1,0,1) scenario and approximate 95% interval for 2025-2030.

#### Supplementary Figure 6 | Observed and age-specific projected C10 death burden.

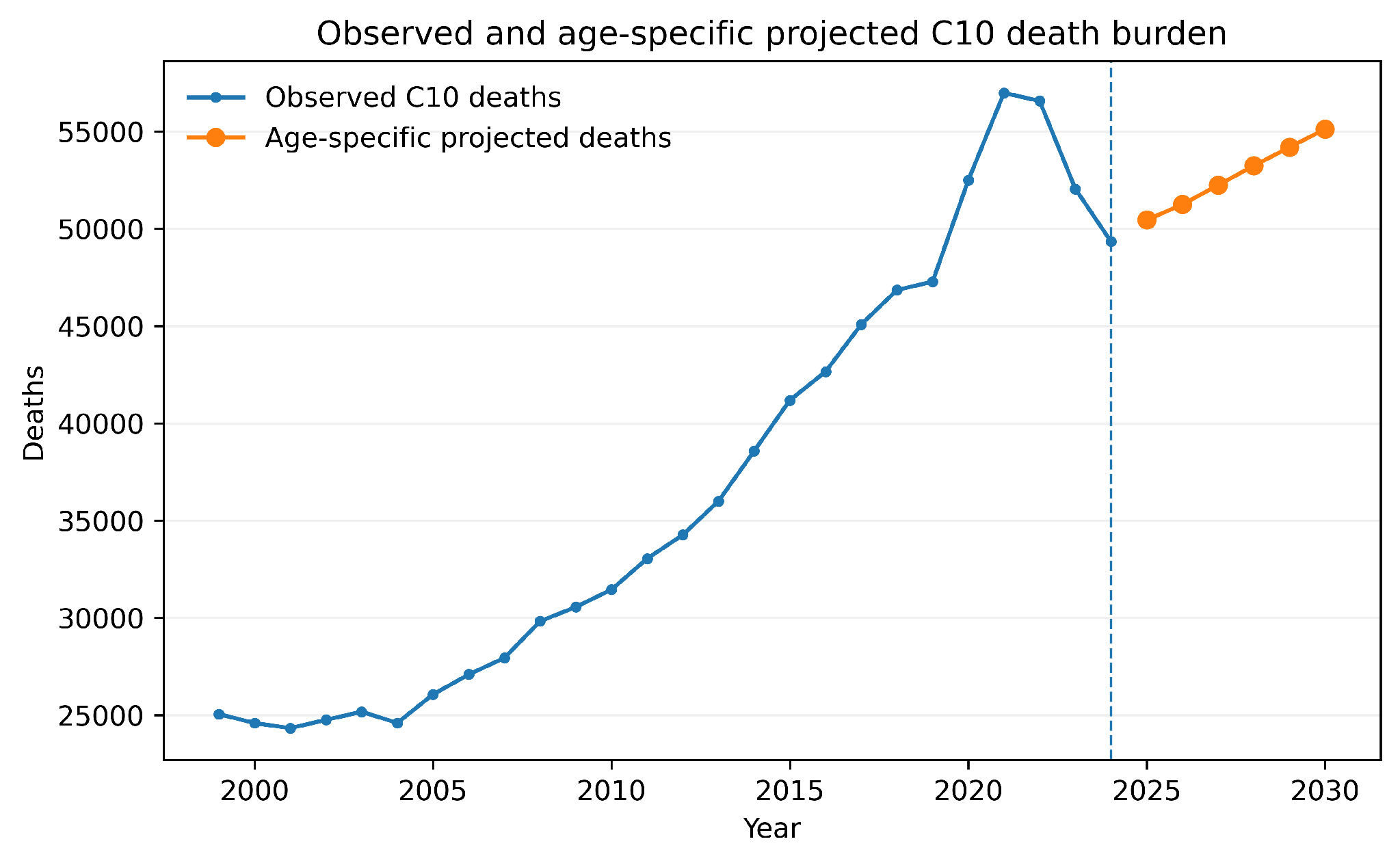

Observed annual C10-associated deaths are shown through 2024, followed by age-specific projected deaths for 2025-2030.
